# Leveraging the CSF proteome toward minimally-invasive diagnostics and biological characterization of brain malignancies

**DOI:** 10.1101/2022.06.17.22276547

**Authors:** Nicholas Mikolajewicz, Shahbaz Khan, Mara Trifoi, Anna Skakdoub, Vladmir Ignatchenko, Sheila Mansouri, Jeffrey Zuccatto, Brad E. Zacharia, Michael Glantz, Gelareh Zadeh, Jason Moffat, Thomas Kislinger, Alireza Mansouri

**Author notes:** Corresponding authors: Alireza Mansouri MD MSc. Department of Neurosurgery, Penn State Cancer Institute, Hershey, PA 17033., Thomas Kislinger PhD. Princess Margaret Cancer Center, University Health Network and Department of Medical Biophysics, University of Toronto, Toronto, Ontario.

## Abstract

1

**Background:** Diagnosis and prognostication of intra-axial brain tumors hinges on invasive brain sampling, which carries risk of morbidity. Minimally invasive sampling of proximal fluids, also known as liquid biopsy, can mitigate this risk.

**Objective:** To identify diagnostic and prognostic cerebrospinal fluid (CSF) proteomic signatures in glioblastoma (GBM), brain metastases (BM), and primary central nervous system lymphoma (CNSL).

**Methods:** CSF samples were retrospectively retrieved from the Penn State Neuroscience Biorepository and profiled using shotgun proteomics. Proteomic signatures were identified using machine learning classifiers and survival analyses.

**Results:** Using 30 µL CSF volumes, we recovered 755 unique proteins across 73 samples. Proteomic-based classifiers identified malignancy with area under the receiver operating characteristic (AUROC) of 0.94 and distinguished between tumor entities with AUROC ≥0.95. More clinically relevant triplex classifiers, comprised of just 3 proteins, distinguished between tumor entities with AUROC of 0.75-0.89. Novel biomarkers were identified, including GAP43, TFF3 and CACNA2D2, and characterized using single-cell RNA sequencing. Survival analyses validated previously implicated prognostic signatures, including blood brain barrier disruption.

**Discussion:** Reliable classification of intra-axial malignancies using low CSF volumes is feasible, allowing for longitudinal tumor surveillance. Based on emerging evidence, upfront implantation of CSF reservoirs in brain tumor patients warrants consideration.

**Statement of Significance:** Current approaches to diagnosing brain tumors risk morbidity. The CSF may be an ideal liquid biopsy matrix for mitigating this risk. We report feasibility of high-throughput CSF proteomics on limited volumes from brain tumor patients with intraventricular reservoirs, demonstrate diagnostic and prognostic utility, and explore its applications in practice.

## 3 Introduction

Advances in the management of brain malignancies have been limited and patients continue to face a grim prognosis.^1–7^ Encompassing a broad category that includes high-grade gliomas, brain metastases, and central nervous system lymphomas (CNSL), brain malignancies pose major challenges in clinical management: *i*) dependence on invasive tumor tissue sampling for initial histopathology-based diagnosis; *ii*) imperfect strategies for tumor surveillance following initial therapy; and *iii*) a lack of clinically actionable, minimally invasive biomarkers.

At presentation, clinical and imaging parameters alone are not always sufficient for definitively distinguishing high-grade gliomas from brain metastases and CNSL. Given the vastly divergent management for each entity, this necessitates direct tissue acquisition through an invasive neurosurgical procedure which presents with great potential for complication. Even minimally-invasive stereotactic brain tumor sampling has a 4-7% risk of major morbidity.^8^ This overt dependence on tumor tissue is an even greater challenge during surveillance while on adjuvant therapy and subsequent treatment. Radiation-based treatment regimens can lead to radiation necrosis (RN) in 10-15% of cases while up to 10% of glioblastoma patients can demonstrate pseudo-progression on imaging.^9, 10^ Despite advances in imaging, pathological tissue assessment remains the gold standard approach for distinguishing RN from true tumor progression. Short interval imaging and clinical follow-up is recommended for differentiating pseudo-progression from true tumor progression. Unfortunately, rapid disease progression is not uncommon, at which point many regimens – including enrolment into clinical trials – are no longer feasible due to advanced disease. While longitudinal sampling of the tumor and its microenvironment is necessary for monitoring the expansion of treatment-resistant subclones or differentiating tumor recurrence from RN, this is simply not feasible for brain-based pathologies. Thus, it behooves us to develop approaches that help avoid unnecessary surgery while tailoring the specific approach based on tumor prognosis when surgery is necessary.

Commonly referred to as liquid biopsy, sampling of proximal fluids has offered valuable insight in various systemic cancers. In Neuro-Oncology, blood and cerebrospinal fluid (CSF) are the most relevant proximal fluids. While acquisition of blood is associated with a theoretically lower risk of morbidity, the CSF is physiologically the ideal liquid biopsy source for brain tumors, owing to its direct contact with the tumor microenvironment in the central nervous system and limited obstruction by the blood-brain barrier. In routine clinical practice, CSF sampling is central in the management of CNSL and has been used for prognostication of medulloblastoma and germ cell tumors.^11^ In GBM and brain metastases, although CSF cytology is used clinically to confirm leptomeningeal spread, molecular analyses are currently not part of routine clinical practice for diagnosis, prognostication, and tumor surveillance.

Numerous liquid biopsy-based studies, utilizing a wide variety of molecular assays, have thus far been conducted to develop better diagnostic and prognostic signatures for GBM.^11–16^ None have been clinically-validated. Circulating tumor DNA (ctDNA), shed predominantly from tumor cell turnover, can be used to genotype GBMs at diagnosis and over the course of the disease. Challenges with ctDNA include its extremely low yield in blood, with only slight improvements detected in the CSF^11^, and limited diagnostic alterations distinguishing normal and cancerous ctDNA.^17^ While providing valuable information, DNA- and RNA-level alterations are unable to predict protein activity, which would be necessary for establishing predictive biomarkers or stratification of patients for use in development of targeted therapeutics.^18^ Recent proteogenomic analyses of GBM tumor tissue have demonstrated incoherence between mRNA and protein expression, suggesting that the proteome serves as a better representation of disease state and underlying biology.^19, 20^

A comprehensive understanding of GBM and brain metastases through liquid biopsy-based proteomic studies would enable a proactive approach to diagnosis, prognostication, and targeted therapy. This would be paradigm-shifting. As such, application of large-scale proteomic approaches in the realm of liquid biopsy are now imperative. In this study, we report the feasibility of high throughput proteomics on limited volumes of CSF samples acquired from patients with brain malignancies, describe its diagnostic and prognostic utility, and explore future applications of this approach in routine clinical practice.

## 4 Results

### 4.1 CSF proteomic can reliably diagnose brain malignancy

We performed unbiased proteomic profiling of 73 CSF samples obtained from patients (mean age 63 years, 43.8% female) diagnosed with normal pressure hydrocephalus (NPH, n = 20), glioblastoma (GBM, n = 22), brain metastasis (BM, n = 17), or primary central nervous system lymphomas (CNSL, n = 14) (**Fig 1**, **Table 1**, **Table S1**). BMs were secondary to non-small cell lung cancer (NSCLC; lung) and invasive ductal carcinomas (breast) in 35.3% and 52.9% of cases, and all primary CNSL samples were diffuse large B-cell lymphomas.

**Figure 1.**
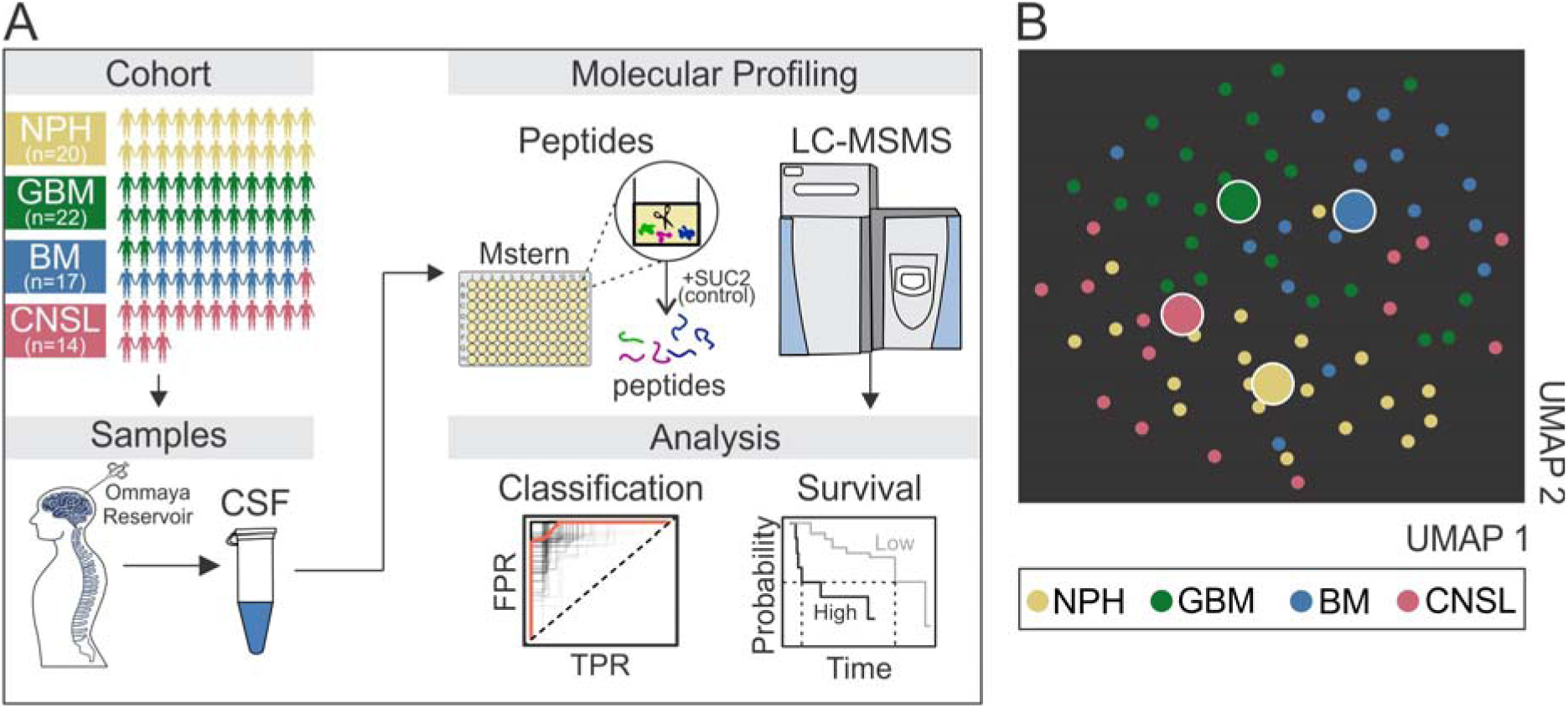
Proteomic characterization of CSF from brain neoplasm patients. **(A)** Schematic representation of study workflow. **(B)** UMAP showing 2D representation of proteomic profiles, with each *small node* representing individual sample and *large nodes* representing median position of each patient cohort. BM; brain metastasis, CNSL; central nervous system lymphoma, FPR; false positive rate, GBM; glioblastoma, LC-MSMS; liquid chromatography with tandem mass spectrometry, NPH; normal pressure hydrocephalus, TPR; true positive rate, UMAP: uniform manifold approximation and projection.

**Table 1.** Patient and sample characteristics.

| **Table 1. Patient and sample characteristics.** |  |  |  |  |  |  |
| --- | --- | --- | --- | --- | --- | --- |
| **Description** | **Statistic** | **NPH** | **GBM** | **BM** | **CNSL\*** | **Total** |
| **Patient Characteristics** |  |  |  |  |  |  |
| **Count** | n | 20 | 22 | 17 | 14 | 73 |
| **Sex** | % ♀ | 40.0 | 27.3 | 71.6 | 42.9 | 43.8 |
| **Age, years** | mean (range) | 73 (48, 91) | 54 (24, 74) | 59 (42, 84) | 68 (57, 86) | 63 (25, 91) |
| **Survival, days** | median | n.d. | 2246 | 564 | 649 | n.d. |
| **Treated** | % | n.a. | 95.5 | 88.2 | 78.6 | 88.7** |
| **CSF Characteristics** |  |  |  |  |  |  |
| **cell count, cells/μL** | mean (sd) | 0 (0) | 18.3 (30.6) | 33.4 (65.9) | 23.6 (57.9) | 25.0 (51.6) |
| **protein content, mg/dL** | mean (sd) | n.d. | 61.7 (56.7) | 70.0 (66.5) | 73.1 (86.5) | 67.5 (67.4)** |
| **malignant cytology** | % positive | n.a. | 19.0 | 70.6 | 25.0 | 38.0** |
| **Tumor characteristics** |  |  |  |  |  |  |
| **tumor burden, cm<sup>3</sup>** | mean (sd) | n.a. | 14.8 (19.4) | 2.4 (4.6) | 19.2 (17.5) | 12.0 (16.8)** |
| **leptomeningeal** | % positive | n.a. | 63.6 | 64.7 | 42.8 | 58.5** |
| **touching ventricle** | % positive | n.a. | 63.6 | 23.5 | 85.7 | 56.6** |
| **IDH status** | % WT | n.a. | 94.1*** | n.a. | n.a. | n.a. |
| **MGMT status** | % methylated | n.a. | 33.3*** | n.a. | n.a. | n.a. |
| **Brain Metastasis Primary Lesion** |  |  |  |  |  |  |
| **lung** | % BMs | n.a. | n.a. | 35.3 | n.a. | n.a. |
| **breast** | % BMs | n.a. | n.a. | 52.9 | n.a. | n.a. |
| **other** | % BMs | n.a. | n.a. | 11.8 | n.a. | n.a. |

\*all CNSL cases are primary CNSL (PCNSL) neoplasms (diffuse large B-cell lymphoma). \*\*only neoplastic samples (GBM, BM and CNSL) included in calculation. \*\*\* IDH and MGMT status determined for 17/22 and 18/22 GBM samples, respectively. Abbreviations: n.a.; not applicable, n.d.; not determined, sd; standard deviation.

For quality control (QC), all samples were spiked with *S. cerevisiae* invertase 2 (SUC2, internal control) and indexed retention time (iRT) peptides (chromatography control). We demonstrated limited variation in SUC2 intensities (**Fig S1A**), and consistent iRT peptide elution profiles, thereby indicating reliable liquid chromatography (LC) performance (**Fig S1B**). To ensure the reliability of our data, comprehensive QC samples were run after a fixed number of biological samples. The pooled QC samples were highly reproducible within processing (R^2^ = 0.92) and technical (R^2^ = 0.93) replicates (**Fig S1C**). Correlation analysis within biological and random groups further confirmed the quality of our data (**Fig S1C**). Using this proteomic workflow, we recovered 1333 unique proteins and showed that CNSL and BM (but not GBM) samples had a significantly more diverse proteome than NPH samples (**Fig S1D**; p = 0.0016 and 0.039, respectively).

Next, we applied a 70% intra-group coverage threshold to obtain a dataset of 755 proteins for downstream analyses, representing an improvement in proteome coverage over the 506 proteins recovered by Schmid and colleagues in a comparable cohort using an identical coverage threshold (**Fig S2A-D**).^16^ Tumor suppressor genes (e.g., PTEN, NF1, TP53) and oncogenes (e.g., EGFR, BRAF, PDGFRA) implicated in GBM, BM and CNSL were not reliably recovered by our protocol, which we attributed to our approach preferentially detecting secreted, rather than intracellular or membrane-bound proteins (**Table S1**).^21^ Importantly, UMAP representation of CSF proteomes revealed coherent separation of each diagnostic group, suggesting that CSF proteomics are suitable for diagnosing malignancy, and discriminating between different brain neoplasms (**Fig 1B**).

To characterize malignancy-associated CSF biology, we performed differential protein analyses between NPH (non-malignant) and each brain neoplasm cohort (malignant) and identified 55 and 112 proteins that were consistently over- or under-represented in malignant CSF samples, respectively (**Fig 2A, B**; **Table S2**). Functional annotation revealed that malignancy was associated with apoptotic signaling, glycolysis and heme metabolism, whereas non-malignancy was associated with elastic fibres- and extracellular matrix (ECM)-associated proteins, and neuronal and glial processes (**Fig 2C, D**; **Fig S3A**). Importantly, using published proteomic CSF data from GBM,^16^ BM,^16^ CNSL,^16^ Alzheimer’s disease (AD),^22^ amyotrophic lateral sclerosis (ALS),^23^ and clinically-isolated syndrome of demyelination [CIS; first attack of multiple sclerosis (MS)],^24^ we demonstrated that our malignancy and non-malignancy signatures are significantly more deregulated in neoplastic disease (GBM, BM, CNSL) than non-neoplastic disease (AD, ALS, CIS/MS) (**Fig 2E, F**).

**Figure 2.**
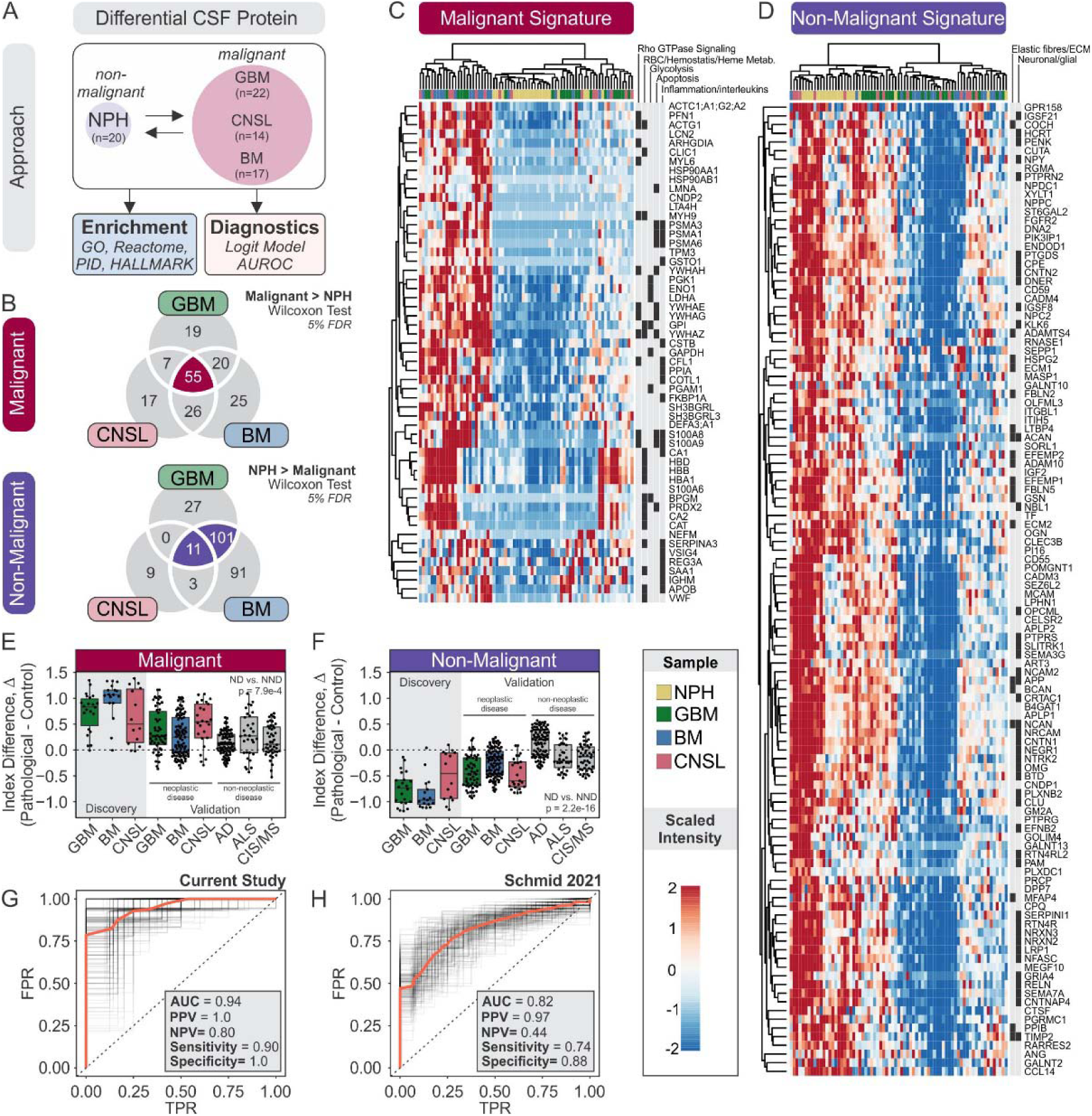
CSF proteomics reliably diagnose malignancy. **(A)** Schematic representation of approach to evaluate value of CSF proteome in classifying malignancy. **(B)** Venn diagram of over- and under-represented protein in brain neoplasms. Malignant and non-malignant protein signatures were derived from the indicated intersections. **(C, D)** Heatmap of scaled intensities of protein associated with malignancy (**C**) and non-malignancy (**D**) signatures. Functional annotations are indicated. **(E, F)** Malignant (**E**) and non-malignant (**F**) index differences (Δ) between disease and control samples. Discovery cohorts include data from current study, and validation cohorts include previously published proteomic data from Schmid et al. (GBM, BM, CNSL)^16^, Bader et al. (AD)^22^, Bereman et al. (ALS)^23^, and Stoop et al. (CIS/MS)^24^. Difference between neoplastic (ND) and non-neoplastic disease (NND) in validation cohorts was assessed by Student’s t-test. **(G)** ROC curve assessing sensitivity and specificity of non-malignant and malignant indices as diagnostic biomarkers in current (discovery) cohort and Schmid (validation) cohort^16^. The indicated performance metrics correspond to *Model 1* in **Table 1**. *Red ROC curve*: median. *Black ROC curves*: each iteration. AD: Alzheimer’s disease, ALS; amyotrophic lateral sclerosis, CIS/MS; clinically-isolated syndrome of demyelination/multiple sclerosis, FPR; false positive rate, ROC; Receiver operating characteristic, TPR; true positive rate.

We utilized a logistic regression (LR) machine-learning classifier framework to evaluate the diagnostic utility of our proteomic signatures. We found that proteomic-based LR classifiers identified malignancy with a median AUROC of 0.94 (95% CI = 0.85 – 1.0), and this estimate ranged from 0.95 – 1.0 when evaluated for single neoplastic entities, demonstrating minimal neoplasm-specific bias (**Fig 2G**, **Table 2**; *Model 1*). These performance metrics were reproduced using L1-regularized LR classifiers (**Table 2**; *Model 2*), and feature selection using the Lasso procedure independently identified SERPINA3, HIST1H1E;HIST1H1D, IGFALS, HBA1, APOC2, FSTL3, SH3BGRL3, FCGR3A, FDPS, HLA-C, IGHM, HYOU1, REG3A, LGALS1, and COL4A1 as malignancy-associated protein, and GALNT2, PI16, AGA, COCH, CCL14, PLXDC1, IGHV1-2 and GNPTG as non-malignancy-associated protein. Despite only 38% and 80% of proteins belonging to the malignant and non-malignant signatures being recovered in the Schmid cohort, respectively (**Fig S3B**),^16^ malignancy in this independent cohort was classified with a median AUROC of 0.82 (95% CI = 0.72 – 0.91), demonstrating the external validity of our signatures (**Fig 2H**, **Fig S3C-H**, **Table 2**).

**Table 2.** Summary of diagnostic classifiers.

| Classifier | Cohort | Data | AUROC (median ± 95% CI) | | Model 1 Metrics<sup>#</sup> | | | | | |
| --- | --- | --- | --- | --- | --- | --- | --- | --- | --- | --- |
|  |  |  | Model 1* | Model 2** | Sens | Spec | PPV | NPV | +LR | -LR |
| Malignancy | Current | train, test | 0.94<br>(0.85,1.0) | 0.92<br>(0.85, 0.99) | 0.90 | 1.0 | 1.0 | 0.80 | Inf | 0.10 |
|  | Schmid | test | 0.82<br>(0.72,0.91) | 0.81<br>(0.75, 0.88) | 0.74 | 0.88 | 0.97 | 0.44 | 6.2 | 0.30 |
| GBM<br>(vs-other) | Current | train, test | 0.95<br>(0.80,1.0) | 0.92<br>(0.82, 1.0) | 0.89 | 1.0 | 1.0 | 1.0 | Inf | 0.11 |
|  | Schmid | test | 0.69<br>(0.56,0.81) | 0.69<br>(0.61, 0.77) | 0.72 | 0.70 | 0.50 | 0.85 | 2.4 | 0.40 |
| BM<br>(vs-other) | Current | train, test | 0.98<br>(0.90,1.0) | 0.98<br>(0.95, 1.0) | 1.0 | 1.0 | 1.0 | 1.0 | Inf | 0.0 |
|  | Schmid | test | 0.71<br>(0.58,0.83) | 0.70<br>(0.62, 0.78) | 0.63 | 0.80 | 0.77 | 0.65 | 3.2 | 0.46 |
| CNSL<br>(vs-other) | Current | train, test | 1.0<br>(0.88,1.0) | 0.97<br>(0.93,1.0) | 1.0 | 1.0 | 1.0 | 1.0 | Inf | 0.0 |
|  | Schmid | test | 0.80<br>(0.64,0.94) | 0.78<br>(0.68,0.89) | 0.80 | 0.80 | 0.40 | 0.96 | 4.0 | 0.25 |

\**Model 1*: LR model. *Resampling*: 70/30 train/test partitions, 200 iterations. Unless otherwise specified, reported results refer to these models to interpret findings in the current study.
\*\**Model 2*: L1-regularized LR model (Lasso). *Resampling*: leave-one-out cross-validation.
<sup>#</sup> Model 1 performance metrics were determined at cutoff at which sensitivity + specificity was maximized.
Abbreviations: +LR; positive likelihood ratio, -LR; negative likelihood ratio, CI; confidence interval, Inf; infinite (resulting from division by zero), LR; logistic regression, n.d.; not determined, NPV; negative predictive value, PPV; positive predictive value, sens; sensitivity, spec; specificity.

Upon consideration of clinical covariates, including age, sex, presence of malignant cytology in CSF, and leptomeningeal status, we observed a negligible 0 - 0.02 improvement in AUROC performance (**Fig S3D, H**). From the clinical covariates considered here, malignant cytology had the best individual diagnostic value with a median AUROC of 0.68 (95% CI = 0.58 – 0.79); however, despite perfect specificity (median = 1.0, 95% CI = 1.0 – 1.0), its poor sensitivity (median = 0.37, 95% CI = 0.16 – 0.58) indicates that additional diagnostic investigations are required following a negative cytological finding. Here we report that our proteomic-based malignancy classifier is suitable for such applications, given its comparable median specificity of 1.0 (95% CI = 0.84 – 1.0) and superior sensitivity of 0.90 (95% CI = 0.73 – 1.0).

### 4.2 Diagnosis of brain neoplasms using CSF proteomics is clinically feasible

We next sought to evaluate whether CSF proteomics can discriminate between different types of brain neoplasms. We approached this question from two perspectives. In the *first*, we evaluated the upper-bound performance of CSF proteomics in diagnosing different types of brain malignancy without consideration for clinical feasibility (i.e., no restriction imposed on number of proteins used to construct classification model) (**Fig S4**; **S5**). However, recognizing that profiling large panels of proteins in a clinical setting may not be readily available in all settings, our *second* approach prioritized clinical feasibility and identified smaller subsets of diagnostic biomarkers in a proof-of-concept demonstration that focused CSF proteomics has diagnostic utility (**Fig 3**; **Fig S6-S7**).

**Figure 3.**
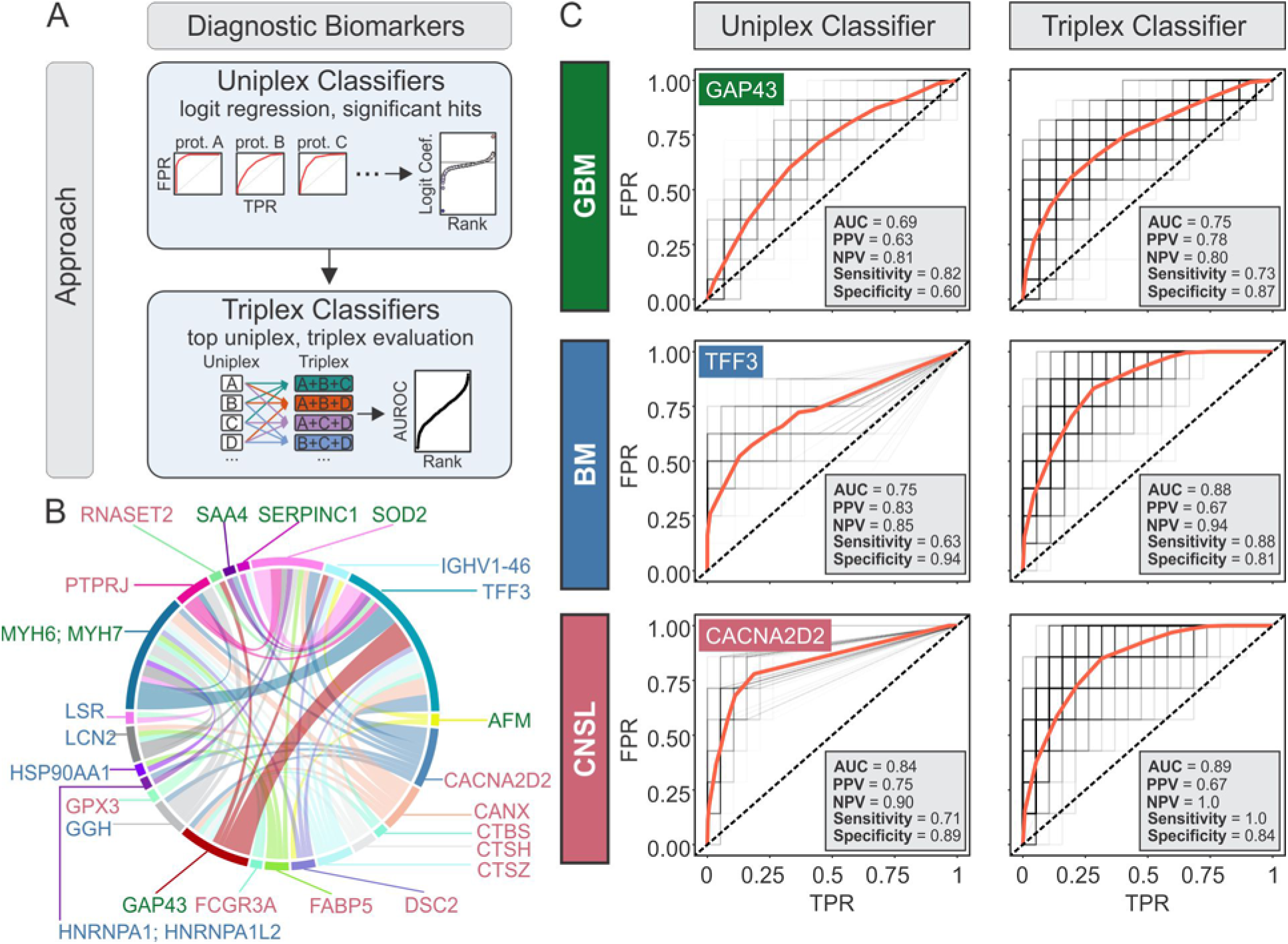
Identification of diagnostic brain neoplasm-specific biomarkers. **(A)** Schematic representation of approach to construct and evaluate uniplex and triplex diagnostic classifiers. **(B)** Relative frequencies of protein co-occurrences among top 25 triplex classifiers. Individual proteins are color coded by neoplasm specificity (*green*: GBM, *blue*: BM, *red*: CNSL). *Circumference of circle* occupied by a protein is proportional to the number of top triplex classifiers utilizing that protein, and *width of cord* connecting two protein is proportional to the number of top triplex classifiers in which both proteins are utilized. **(C)** Comparison of ROC curves for uniplex and triplex classifiers, using GAP43, TFF3 and CACNA2D2. Uniplex classifiers use one protein as a predictor, whereas triplex classifiers use all three proteins as predictors. Performance metrics indicated in plots correspond to *Model 1* in **Table 3**. *Red ROC curve*: median. *Black ROC curves*: each iteration. FPR; false positive rate, TPR; true positive rate.

**Table 3.** Performance of uniplex and triplex classifiers using GAP43, TFF3 and CACNA2D2

| Classifier | Predictors | AUROC<br>(median ± 95% CI) | | Model 1 Metrics<sup>#</sup> | | | | | |
| --- | --- | --- | --- | --- | --- | --- | --- | --- | --- |
|  |  | Model 1* | Model 2** | Sens | Spec | PPV | NPV | +LR | -LR |
| GBM<sup>a</sup><br>(vs-other) | GAP43 | 0.69<br>(0.55, 0.84) | n.d. | 0.82 | 0.60 | 0.63 | 0.81 | 2.1 | 0.30 |
| BM<sup>a</sup><br>(vs- other) | TFF3 | 0.75<br>(0.58, 0.92) | n.d. | 0.63 | 0.94 | 0.83 | 0.85 | 11.0 | 0.39 |
| CNSL<sup>a</sup><br>(vs- other) | CACNA2D2 | 0.84<br>(0.72, 0.96) | n.d. | 0.71 | 0.89 | 0.75 | 0.90 | 6.5 | 0.33 |
| GBM<sup>b</sup><br>(vs- other) | GAP43 + TFF3 +<br>CACNA2D2 | 0.75<br>(0.61, 0.89) | 0.74<br>(0.59, 0.88) | 0.73 | 0.87 | 0.78 | 0.80 | 5.6 | 0.31 |
| BM<sup>b</sup><br>(vs- other) | GAP43 + TFF3 +<br>CACNA2D2 | 0.88<br>(0.74, 1.0) | 0.88<br>(0.79, 0.97) | 0.88 | 0.81 | 0.67 | 0.94 | 4.6 | 0.15 |
| CNSL<sup>b</sup><br>(vs- other) | GAP43 + TFF3 +<br>CACNA2D2 | 0.89<br>(0.72, 1.0) | 0.85<br>(0.75, 0.95) | 1.0 | 0.84 | 0.67 | 1.0 | 6.3 | 0.0 |

\*Model 1: LR model. *Resampling*: 70/30 train/test partitions, 200 iterations. Unless otherwise specified, reported results refer to these models to interpret findings in the current study.
\*\*Model 2: L1-regularized LR model (Lasso). *Resampling*: leave-one-out cross-validation.
<sup>#</sup> Model 1 performance metrics were determined at cutoff at which sensitivity + specificity was maximized.
<sup>a</sup>uniplex classifier.
<sup>b</sup>triplex classifier.
Abbreviations: +LR; positive likelihood ratio, -LR; negative likelihood ratio, CI; confidence interval, LR; logistic regression, n.d.; not determined, NPV; negative predictive value, PPV; positive predictive value, sens; sensitivity, spec; specificity.

To evaluate the upper-bound performance of CSF proteomics in diagnosing different brain neoplasms, we performed pairwise differential protein analyses to identify GBM-, BM- and CNSL-specific proteins and used these to construct neoplasm-vs-other classifiers (**Fig S4A**, **Table S2**). Using the current study’s cohort, the median AUROC for GBM-vs-other, BM-vs-other, and CNSL-vs-other LR classifiers was 0.95 (95% CI = 0.80 – 1.0), 0.98 (95% CI = 0.90 – 1.0), and 1.0 (95% CI = 0.88 – 1.0) (**Fig S4B-D**, **Table 2**; *Model 1*), respectively, and these performance metrics were independently verified using a L1-regularized LR classifier (**Table 2**; *Model 2*). We also retrained this model using the subset of IDH-wt GBM samples (16/22 glioma samples) and found that IDH-wt GBM was classified with a similar AUROC of 0.97 (95% = 0.88 – 1.0). Next, we assessed external validity using data from the Schmid cohort, and found the performance of the GBM-vs-other, BM-vs-other, and CNSL-vs-other LR classifiers to be 0.69 (95% CI = 0.56 – 0.81), 0.71 (95% CI = 0.58 – 0.83), and 0.80 (95% CI = 0.64-0.94), respectively (**Fig S5, Table 2**; *Model 1*). While the performance was lower than that observed in our own cohort, this was attributed to the Schmid cohort recovering only 63% (107/170) proteins used in training the classifier (**Fig S5A**). We conclude that our proteome-based classifiers are generalizable and can discriminate between different neoplastic diseases. However, given that this classifier requires profiling of 170 protein, its utility in a clinical setting is limited, with the exception of large-scale targeted proteomic approaches [e.g., parallel reaction monitoring (PRM)].^25–27^

Having established that the upper-bound AUROC performance of CSF proteomics ranges from 0.95 - 1.0, we next sought to identify focused biomarker panels. We applied our machine-learning-based framework to nominate individual protein with possible diagnostic value (**Fig 3A**; **Fig S6A**; **Table S3**). Using an orthogonal approach, we also identified candidate biomarkers using Lasso-based feature selection but found this list of proteins to be redundant with the approach described above. Next, we evaluated combinatorial classifiers, termed “triplex classifiers”, that comprised of 3-way combinations of the top proteins identified by our uniplex classifiers, and these classifiers were AUROC-ranked to identify the top combinations of diagnostic biomarkers (**Fig 3A**; **Fig S6B**; **Table S3**). Among the top 25 triplex classifiers, we focused on further characterizing GAP43 (GBM-specific), TFF3 (BM-specific), and CACNA2D2 (CNSL-specific) (**Fig 3B**). Whereas GAP43, TFF3 and CACNAD2 uniplex classifiers had a median AUROC of 0.69, 0.75 and 0.84, in classifying GBM, BM and CNSL, respectively, the combined triplex classifier performed better with a median AUROC of 0.75, 0.88 and 0.89, respectively (**Fig 3C**, **Table 3**). We also evaluated these uniplex and triplex classifiers on the subset of IDH-wt GBM (16/22 glioma samples) and demonstrated AUROCs of 0.68 (95% CI: 0.465 – 0.87, p = 0.04) and 0.75 (95% CI: 0.58 - 0.95, p = 0.01), respectively.

To explore the biology of these biomarkers, we assessed their transcriptional profiles using publicly-available single cell transcriptomic (scRNAseq) profiles of GBM,^28^ lung adenocarcinoma BM,^29^ and CNSL (**Fig S7**).^30^ In GBM scRNAseq data, GAP43 expression was largely restricted to GBM cells, limited in lymphoid and myeloid cells, and entirely absent in BM and CNSL cells. TFF3 was exclusively expressed in BM. CACNA2D2 expression was absent in GBM and CNSL and detected in a subpopulation of BM tumor cells. Collectively these data suggest that GAP43 and TFF3 observed in CSF are derived from tumor populations, unlike other candidate biomarkers (e.g., SOD2, PZP, and CTSZ) that exhibit non-specific patterns of expression. The lack of transcriptomic CACNA2D2 expression in CNSL samples suggests a non-neoplastic source, or lack of correlation between transcriptomic and proteomic expression.

### 4.3 Pathway-level analyses identify survival associations

In our cohort of brain tumor patients, BM and CNSL patients had similar survival rates (564 and 649 median days, respectively), whereas GBM patients had a median survival of 2246 days (**Fig 4A**). To explore the biology associated with survival, we evaluated seven diverse cancer-associated signature sets in our cohort, including pan-cancer and GBM-specific signatures from the Clinical Proteomic Tumor Analysis Consortium (CTPAC).^20, 31–36^ In addition to these signatures discriminating malignant from non-malignant samples (**Fig S8A**), survival analyses revealed that mesenchymal and invasion-associated signatures trended towards unfavorable survival outcomes (**Fig S8B**). Our small sample size limited us from identifying novel prognostic biomarkers (**Fig S9A-D**, **Fig S10A**). To overcome this limitation, we undertook a pathway-level analysis which validated previous findings reported by Schmid et al. (**Fig 4B-D**, **Fig S9E**, **Fig S10B-E**).^16^ Specifically, we demonstrated that markers associated with blood-brain barrier disruption [e.g., blood coagulation (C4BPB, COL1A1, CPB2, F10, F13B, F2, F9, FGA, FGG, PLG, PROS1, PROZ, SERPINA1, SERPINA5, SERPINC1, SERPIND1, VWF), and complement pathway activation (C2, C3, C4B, C4BPA, C4BPB, C5, C8A, C8B, C9, CFB, CFH, CFHR1, CFI, CPB2, CPN1, CPN2, F2, SERPING1, VTN)] (**Fig 4B**), angiogenesis (APOE, APOH, CDH5, HRG, PROC, SEMA6A, THBS1) (**Fig 4C**) and stemness (KIT, NOTCH2) (**Fig 4D**) were associated with unfavorable survival outcomes.

**Figure 4.**
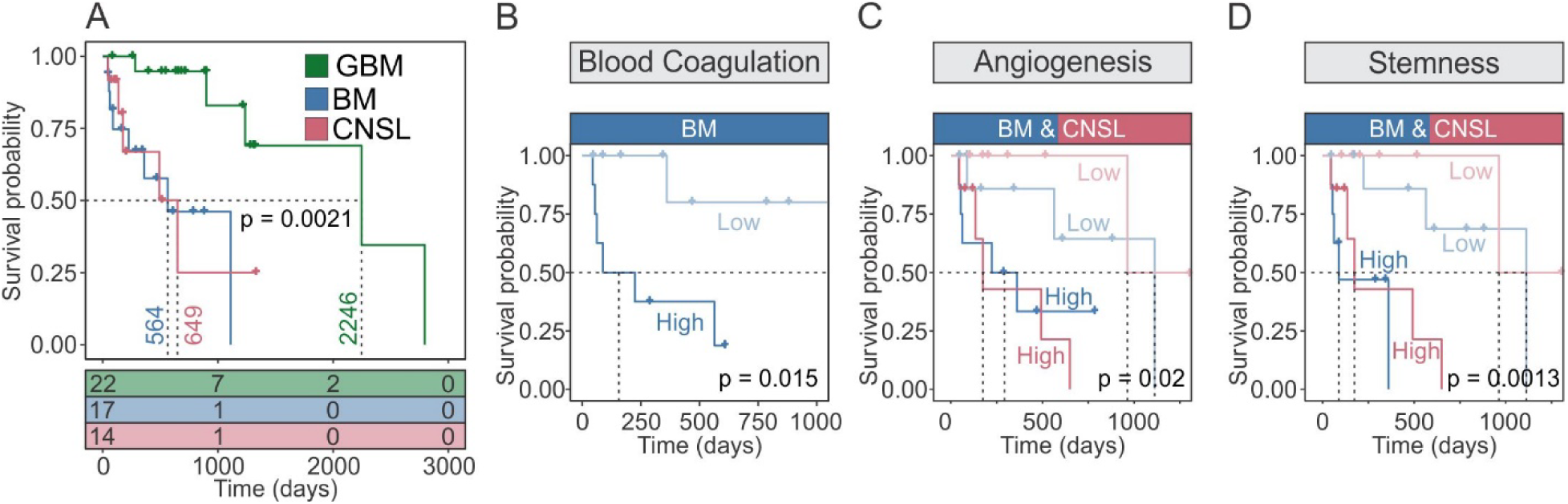
Survival analyses. **(A)** Kaplan Meier survival curve of patient cohort stratified by brain neoplasm diagnosis. **(B-C)** Kaplan Meier survival curves showing patient survival between pathway-stratified groups (high vs. low; split at median) using blood coagulation- (**B**), angiogenesis- (**C**), and stemness- (**D**) associated proteins. BM; brain metastasis, GBM; glioblastoma, NPH; normal pressure hydrocephalus, CNSL; central nervous system lymphoma.

## 5 Discussion

In this proof-of-concept study, we show the feasibility of high protein recovery and comprehensive proteomic analyses on low sample volumes, using the MStern sample processing approach.^37^ We recovered 755 proteins with as little as 30 µL of CSF per sample. Along with clearly distinguishing malignant from non-malignant samples, we were able to build and externally validate classifiers for distinguishing GBM, BM, and CNSL. With an eye toward relevance to clinical practice, we further developed a triplex classifier comprised of GAP43, TFF3, and CACNA2D2 that can together distinguish between the three disease entities. Prospective validation of these findings can have a profound impact on the diagnosis and longitudinal surveillance of these malignant entities.

Using CSF based proteomics, we were able to distinguish malignant from non-malignant samples and identified pathway-level alterations of prognostic significance. These were both externally validated using data from the Schmid et al. cohort.^16^ While CSF proteomics offered similar diagnostic specificity to CSF cytology alone, we demonstrated that CSF proteomics outperformed CSF cytology with respect to sensitivity, thereby representing a significant value-add for distinguishing malignant from non-malignant samples. We were also able to develop machine learning based classifiers to reliably distinguish between the three brain tumor entities; this was again externally validated using the Schmid cohort.^16^ Since these classifiers depended on larger protein panels, traditional antibody-driven methods, such as Western Blotting and ELISA, are inefficient and represent a potential bottleneck. Selective reaction monitoring (SRM) and, more recent, parallel reaction monitoring (PRM) utilize stable isotope-labeled peptides as internal standards of previously detected candidates. Targeted proteomics assays are quantitative tools that enable the robust, sensitive and multiplexable quantitation of proteins without the need of antibodies,^38, 39^ but require significant time for initial development, validation and implementation of these assays. Thus, recognizing that large-scale mass spectrometric analyses can be challenging in the routine clinical setting, we further built on our approach by identifying the top 3 biomarker proteins that can together distinguish between GBM, BM, and CNSL with high precision (AUROC 0.75–0.89). Further prospective validation of these biomarkers will be necessary.

Beyond simply distinguishing between the three tumor entities, the identification of GAP43, TFF3, and CACNA2D2 as biomarkers specifically associated with GBM, BM, and CNSL, respectively, can have value for longitudinal monitoring of patient during adjuvant therapy and surveillance as well. Specifically, in the case of GAP43 and GBM, we were able to show that the expression of the protein is largely restricted to tumor cells, with little expression in lymphoid and myeloid cells. This has significant implications in monitoring for true tumor recurrence and distinguishing it from other common pathological entities, such as RN. This further supports the need to consider upfront implantation of CSF reservoirs for routine monitoring of the tumor microenvironment during therapy and surveillance, through scheduled sampling of the CSF to assess for relevant biomarkers. Currently, such strategies have rarely been implemented. In 2016, Brown et al. used CSF reservoirs for the delivery of CART cell therapy, with regular monitoring of cytokines and immune cells within the CSF, in a patient with multifocal recurrence of GBM with remarkable success.^40^ This was part of a Phase 1 trial that is currently underway (<u>NCT02208362</u>).

The Trefoil Factor Family (TFF1, TFF2, and TFF3) are proteins secreted by normal mucous secretory epithelia.^41^ TFF3 transcription and translation has been reported in various cancers. The oncogenic behavior of TFF3 is mediated through promotion of cancer cell migration and invasion, along with down regulation of apoptotic signaling.^42–44^ In breast cancer, its expression has been associated with lymph node involvement and increased propensity for local metastasis. Of note, its mRNA expression has been linked to higher likelihood of breast cancer metastases to the CSF and bones.^45–47^ Other cancers in which TFF3 has been associated with include gastric,^48^ pancreatic,^49^ colorectal,^50^ cervical,^51^ and prostate.^52^ As such, detection of TFF3 within the CSF can serve as a pan-cancer biomarker of metastases to the CSF. In addition to their role in diagnosis and surveillance following treatment, such biomarkers can potentially be used to screen for CNS involvement in patients with cancer types that are known to have a high propensity for CNS metastases, such as advanced non-small cell lung cancer, or triple negative and HER2+ breast cancer.^53–55^

We found a significant association between CNSL and CACNA2D2 expression, a voltage-gated calcium channel receptor found in numerous tissues including the brain.^56, 57^ The role of this protein in cancer is unclear, with conflicting data on whether it is a tumor suppressor^58^ or oncogenic protein.^59^ While CACNA2D2 could have simply been shed from normal brain tissue, its specific association with CNSL warrants exploration. This is particularly relevant given the emerging role of extracellular vesicles (EVs), including exosomes, in cancer biology. EVs are cell-derived vesicles released by all cells,^60^ and their protective lipid membranes permits protected travel throughout the CSF and blood stream in the body.^61^ Complex interactions by cancer cells with other cells in the tumor microenvironment can be mediated through EVs, via exchange of biologic factors including DNA, RNA, and proteins.

### Limitations

Our findings should be interpreted with several caveats, including the retrospective nature of the study precluding comprehensive correlative analyses with clinical data. Furthermore, given the retrospective acquisition of samples, certain sample characteristics were unavailable, including IDH status in 5/22 glioma samples. Similarly, most gliomas included in this study were classified prior to the updated 2021 WHO guidelines, thus it is possible that some glioma samples in which IDH status was not determined are not grade IV GBM. Within the BM cohort, it was not documented whether CSF was acquired while the primary tumor still existed or not, and this could represent a potential confounding variable. The exquisitely long survival of our GBM cohort (median of 2246 days), a selection of ultra-long survivors by virtue of our clinical protocol for implantation of CSF reservoirs in GBM patients, also necessitates the cautious interpretation of our data pertaining to this tumor entity. Although our triplex classifier presents the opportunity for a clinically facile approach to reliably distinguishing between the three cancer entities and potentially surveillance for tumor recurrence, neither of GAP43, TFF3 or CACNA2D2 were recovered in the Schmid cohort,^16^ thereby precluding evaluation of their external validity in an independent cohort. Although corroboration with tumor-derived scRNAseq data offered exciting insight into the potential source of proteins detected in our CSF samples, it must be emphasized that scRNAseq data were derived from tumor tissue and not CSF samples. Similarly, signatures and associations identified in this study are restricted to free proteins shed into the CSF and do not reflect a comprehensive snapshot of the composition of the tumor microenvironment. Our current analysis does not account for alternate splice site variations and post-translational modifications. In addition to correlative analyses with matched tumor and plasma, analyses of exosome cargo and incorporation of other -omic analyses, including phosphoproteomics, glycoproteomics, metabolomics, genomics, and lipidomics will be critical as well.^20^ Despite these limitations, our study is the first of its kind to demonstrate the feasibility of high protein recovery from very limited CSF sample, yielding externally validated diagnostic signatures that can readily be applied in the clinical setting.

### Future Directions

Given the retrospective nature of the current study, prospective validation of the proteomic signatures identified will be required. In parallel, development and optimization ELISA and PRM-based assays for small- (e.g., triplex classifiers) and large-panel protein signatures, respectively, will facilitate the translation of the diagnostic classifiers into a clinical setting. In particular, PRM enables targeted quantification of tens to hundreds of proteins, and while is primarily used as a research tool, its scalability, cost-effectiveness, and information-rich readouts make it a promising tool for future clinical applications. Finally, while our glioma cohort was predominantly comprised of IDH-wt GBM patients, we recognize the value of using a CSF proteomic approach to discriminate between different glioma entities, including IDH-mutant gliomas, and future studies addressing this will be required.

## 6 Materials and Methods

The current study adhered to the Reporting Recommendations for Tumor Marker Prognostic Studies (REMARK) guidelines. The completed checklist is provided in **Appendix 1**.

### Software

Figure preparation: CorelDRAW x8 (Corel); Bioinformatic analyses: R version 4.0.3 (R Foundation for Statistical Computing).

### Patient Cohort

#### Study eligibility

All patients with a diagnosis of a brain tumor under the care of a physician within the Penn State Hershey Neuroscience Institute who provided informed consented for the Biorepository study, were eligible.

#### Patient recruitment

Any patient potentially eligible for the Penn State Hershey Neuroscience Institute Biorepository was first identified by the physician responsible for their care. The physician informed the research coordinator who then independently explained the study to the patient and obtained informed consent. Specimens were only acquired as residuals from samples being collected for routine clinical care or necessary surgical procedures.

#### Ethics

The Biorepository study is approved by the Penn State IRB (#2914). Specimens and associated data were released to the protocol “Molecular testing of nervous system cancers as a classification tool” following approval of the Biorepository Data Oversight Committee (#20-0002). Data was released in a de-identified manner via an honest broker system. The patient’s privacy was protected in accordance with both Penn State’s IRB and HIPAA guidelines.

### Proteomic Profiling

#### Sample acquisition

Most CSF samples were collected from intracranially-implanted CSF reservoirs. For lumbar puncture-acquired samples, an atraumatic 21-gauge spinal needle was used. The volume withdrawn ranged from 5-15 mL per collection. Samples were aliquoted into 1 mL polypropylene tubes and all samples were processing within 0-4 hours after collection, during which the samples were stored at room temperature. The tubes were then spun at 2000 g for 10 min at room temperature to remove cellular debris. Supernatants were maintained in 1 mL aliquots and stored at -80 °C. Although higher volumes were available, we only required 30 µL of CSF for reliable proteomics results. In addition, QC samples were created by mixing an equal volume of all 73 sample. Five QC samples were prepared separately, and each QC was run in technical duplicates on the instrument to monitor technical variabilities.

#### Proteomics

Protein concentrations were determined by BCA assay (Pierce) and a volume equivalent to 25 µg of protein was used for sample processing, and each sample was spiked in with 2 pmol of yeast invertase (SUC2) as a sample processing control. The samples were denatured and alkylated with DTT and iodoacetamide, respectively. CSF proteins were purified using an adapted MStern technique.^62^ The samples were bound to a PVDF 96-well MStern plate (Millipore) facilitated by a vacuum suction manifold (Millipore). Adsorbed proteins were washed with 100 mM ammonium bicarbonate (pH = 8) and digested for 4 h at 37 °C via the addition of 50 µL of digestion buffer (5% acetonitrile, 100 mM ABC, 1 mM CaCl_2_) containing 2 µg of trypsin-LysC protease mixture (Promega). The resultant peptides were eluted from the membranes with 50% acetonitrile, lyophilized and desalted with C18 solid-phase extraction tips prepared in-house. 10 µL of purified peptides was spiked with 1 µL of indexed retention time (iRT) (Biognosys) peptide standard. Overall, 11 µL of peptides were loaded onto a 2 cm PepMap Acclaim trap column (Thermo Scientific) using an Easy1000 nanoLC (Thermo Scientific). The peptides were separated and detected along a two-hour reversed-phase gradient using a 50cm EasySpray analytical C18 column coupled by electrospray ionization to a Q-Exactive HF Orbitrap mass spectrometer (Thermo Scientific) operating in a Top 20 data-dependent acquisition mode. MS^1^ data was acquired at a resolution of 120,000 with an AGC target of 1e6 ions and a maximum fill time of 40 ms. MS^2^ data was acquired at a resolution of 30,000 with an AGC target of 2e5 ions and a maximum fill time of 55 ms. A dynamic exclusion of 20 s was enabled, the S-lens RF was set to 59% and the normalized collision energy was set to 27%. The acquired raw data was searched using Maxquant (version 1.6.3.3) against a UniProt complete human protein sequence database (v2020_05) also including yeast invertase (SUC2) and iRT standard peptides.^63^ Two missed cleavages were permitted along with the fixed carbamidomethyl modification of cysteines, the variable oxidation of methionine and variable acetylation of the protein N-terminus. Relative label-free protein quantitation was calculated using MS1-level peak integration along with the matching-between-runs feature enabling a 2 min retention time matching window. False discovery rate (FDR) was set to 1% for peptide spectral matches and protein identification using a target-decoy strategy. The protein groups file was filtered for proteins detected by a minimum of two peptides and then used to carry out further analysis. Missing LFQ values were imputed with normalized iBAQ intensities.^64^

### Bioinformatic Analyses

#### Data sources

Proteomic data from Schmid et al. was obtained from ProteomeXchange (ID: <u>PXD021984</u>);^16^ Bader et al. from ProteomeXchange (ID: <u>PXD016278</u>);^22^ Bereman et al. from Dataset 1 file of electronic supplementary material of original publication;^23^ and Stoop et al. from Supporting Information File 2 of original publication.^24^ Single cell RNA seq data from Wei et al. (2021) was obtained from Gene Expression Omnibus (GEO; accession number <u>GSE181304</u>);^30^ Neftel et al. (2019) from GEO (accession number <u>GSE131928</u>);^28^ and Kim et al. (2020) from GEO (accession number <u>GSE131907</u>).^29^

#### Data preprocessing

Protein X patient intensity matrices were loaded into a Seurat object (*Seurat* 4.0.4 R package^65–68^) and sample normalization (column-wise) and protein scaling (row-wise) of log-transformed data was performed. Protein identified in at least 70% of samples within a diagnostic group were retained for downstream analysis.^16^ No imputations for missing values were performed. For each sample, meta data included a patient identifier, clinical diagnosis (NPH, GBM, BM, CNSL), age, sex, CSF cytology (presence of malignant cells), and leptomeningeal status, alive status and survival time. Data from the Schmid cohort were processed identically, however the only meta data that were available were clinical diagnosis, age and sex.^16^

#### Uniform manifold approximation and projection

To generate a two-dimensional representation of CSF proteomes, the scaled protein X patient intensity matrix was dimensionally-reduced using robust principal component analysis [*PcaHubert*(…, k = 50, kmax = 50, maxdir = 100, signflip = T), *rrcov* v1.6-0 R package^69^] and the top 30 principal components were used for UMAP embedding [*RunUMAP*(…, dims = 1:30), *Seurat* package].

#### Differential protein analysis

Differentially-expressed proteins between two groups were identified by two-sided unpaired Wilcoxon test (*wilcoxauc* function, *presto* v1.0.0 R package^70^). To ensure results were robust to outliers, we used a resampling procedure that involved 100 iterations of differential expression analysis performed on 95% randomly sampled subsets of data. Protein that were significant at 5% false discovery rate (FDR; Benjamini-Hochberg method implemented using *p.adjust* function, *stats* v4.0.3 R package) across all iterations were used in downstream analyses.

#### Functional annotation

Protein signatures were functionally annotated by performing hypergeometric overrepresentation analysis (*fora* function, *fgsea* v1.14.0 R package^71^) using Gene Ontology (GO) biological processes (BP) and cellular components (CC), protein interaction database (PID), HALLMARK gene sets^72^, and Reactome^73^ databases. Enrichments were ranked by Benjamini-Hochberg-adjusted p-values (q-value), and the top 5 annotations for each database were shown.

#### Classification models

For LR classification models (**Table 2**; *Model 1*, **Table 3**; *Model 1*), training and testing cohorts were generated by randomly splitting CSF samples into 70% and 30% subsets, respectively. The training set was used to train a binomial generalized linear model (GLM) using *bayesglm*(…, family = ‘binomial’, maxit = 500) (*arm* R package, v1.12-2^74^) and the AUROC performance was evaluated in the test set (*performance* function, *ROCR* v1.1-11 R package).^75^ Given our smaller cohort size, we confirmed that performance were not influenced by choice of partition ratio (50:50, 60:40 and 80:20 train:test splits yielded similar results). This resampling procedure was repeated over 200 iterations to obtain stable estimates of the median AUROC and corresponding 95% confidence intervals. ROC curves for each iteration were generated using results from *performance*(…, measure = ‘tpr’, x.measure = ‘fpr’), and the median ROC curve summarizing overall performance was computed by aggregating the true positive rate (TPR) and false positive rate (FPR) estimates across all iterations. For each model, sensitivity, specificity, positive predictive value (PPV) and negative predictive value (NPV) estimates were obtained using cut-offs at which TPR + (1-FPR) were maximized. When indicated (**Fig S3F-H**), classifiers were also adjusted for leptomeningeal status, presence of malignant cytology in CSF, sex, and age.

For protein signature-based classifiers, (**Fig 2**, **Fig S3-S5**, *Models 1* and *2* in **Table 2**), signatures were aggregated separately for train and test sets using *gsva*(…, method = “gsva”) (*GSVA* R package, v1.36.2^76^). Given our limited sample size and goal to also characterize biology associated with malignancy, signature discovery (i.e., differential expression analyses, described above) was performed on the total cohort (not training cohort). While in certain contexts such an approach risks leaking information between training and testing cohorts, the resampling procedure used in deriving each signature was tailored to minimize cohort-specific biases. Furthermore, in addition to evaluating the performance of each model on the test cohort derived from the current study’s patient samples, we evaluated the external validity of each trained model on matched samples from the entire Schmid et al. cohort.^16^

For uniplex (single-protein) and triplex (three-protein) classifiers (**Fig 3**, **Fig S5**), protein intensities were not aggregated prior to training the model, and instead intensity values were used as model inputs. For triplex classifiers, the top performing protein combinations were rank-ordered by the average AUROC across GBM-vs-other, BM-vs-other, and CNSL-vs-other classifiers.

To ensure the robustness of the LR models described above, we independently evaluated L1-regularized (i.e., Lasso) LR models (*glmnet* function, *glmnet* R package v1.36.2) using the leave-one-out cross-validation procedure implemented in caret (*caret* R package v6.0-88). For these models, AUROC estimates were calculated and 95% CI were computed using 2000 bootstrap replicates (*ci.auc* function, *pROC* R package v1.18.0) (**Table 2**; *Model 2*, **Table 3**; *Model 2*).

#### Survival analysis

The prognostic value of individual protein was evaluated using univariate Cox proportional hazards regression models (*coxph* function, *survival* v.2-13 R package). The resulting hazard ratios (HR) were visualized on a volcano plot, with the top hits indicated. Given our limited statistical power for survival analyses, p-values with no multiple testing corrections were reported to highlight the strongest associations with survival. Survival-associated pathways were determined by performing gene set enrichment analysis (GSEA; *fgsea* function, *fgsea* v1.14.0 R package^71^) on HR-ranked proteins. GSEA plots were generated using the *plotEnrichment* function (*fgsea* R package). Kaplan Meier survival curves showing patient survival between pathway-stratified groups (high vs. low; split at median) were generated by aggregating protein signature scores with *gsva*(…, method = “gsva”) (*GSVA* R package, v1.36.2)^76^ and visualizing survival with *ggsurvplot*(…) (*survminer* R package, v0.4.9).^77^

#### Single cell transcriptomic analysis

ScRNAseq data sets were normalized, scaled, dimensionally-reduced and visualized on a UMAP using the *Seurat* (version 4.0.4) workflow.^65–68^ In brief, count matrices were loaded into a Seurat object and normalized using *NormalizeData*(…, normalization.method = ‘LogNormalize’, scale.factor = 10000). Variable features were identified using *FindVariableFeatures*(…, selection.method = ‘mvp’, mean.cutoff = c(0.1,8), dispersion.cutoff = c(1,Inf)) and then data were scaled using *ScaleData*(…). Principal component analysis and UMAP embedding was performed using *RunPCA*(…) and *RunUMAP*(…, dims = 1:30), respectively. Metadata from original publications were used to annotate cell types, and gene expression was visualized on a UMAP using *FeaturePlot*(…).

#### Data visualization

Unless otherwise specified, the *ggplot2* R package (version 3.3.5) was used for data visualization. Venn diagrams were generated using either *ssvFeatureEuler* (*seqsetvis* R package, version 1.8.0) or *ggVennDiagram* (*ggVennDiagram* R package, version 1.1.4). Heatmaps were generated using *pheatmap* (*pheatmap* R package, version 1.0.12).

#### Data availability

Proteomic data have been deposited to <u>MassIVE</u> (identifier: MSV000089062). The corresponding FTP link is ftp://massive.ucsd.edu/MSV000089062/.

## Data Availability

All data produced in the present study are available upon reasonable request to the authors

ftp://massive.ucsd.edu/MSV000089062/.

## 8 Author Contributions

Study conception and design: TK, AM

Acquisition of experimental data: MT, AS, SK, MG, TK, AM

Analysis and interpretation of data: NM, SK, TK, AM

Drafting of Manuscript: NM, SK, AM

Supervision: TK, AM

All authors contributed to the critical revision and approval of the final manuscript.

## 9 Acknowledgements

This research work was supported by the 2020 William Donald Nash Brain Tumour Research Fellowship awarded to NM. This work was partially supported by a CIHR Project Grant (PJT154357) to TK. We thank Andrew Macklin for technical assistance. This research was funded in part by the Ontario Ministry of Health and Long-Term Care. TK was supported through the Canadian Research Chair program.

## 10 Tables

Table S1. Protein intensities and clinical characteristics.

Table S2. Protein Signatures.

Table S3. Uniplex and triplex classifier model results.

## 12 Supplemental Figure Legends

**Figure Supplement 1.**
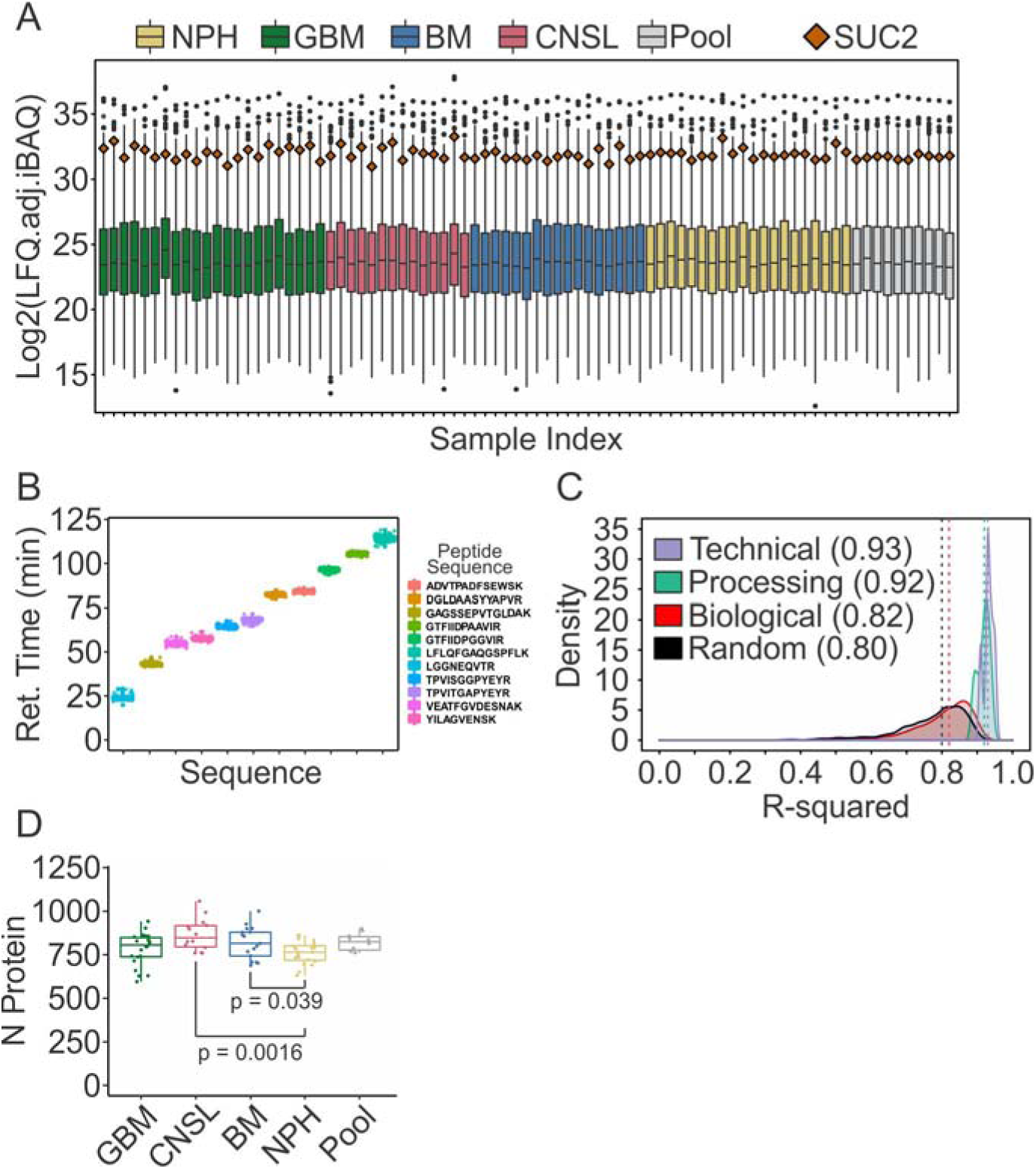
Quality control. **(A)** Boxplot showing log_2_ intensity per sample, with SUC2 intensities shown in in each sample. **(B)**. Average retention time of 11 iRT peptides across each sample with a 135 mins LC gradient. **(C)** Density plot showing median R^2^ correlation between pooled QC samples, run as technical and processing duplicates. The median R^2^ correlations within each of the biological group and within the random groups are also shown. **(D)** Boxplot showing the number of proteins quantified in each group.

**Figure Supplement 2.**
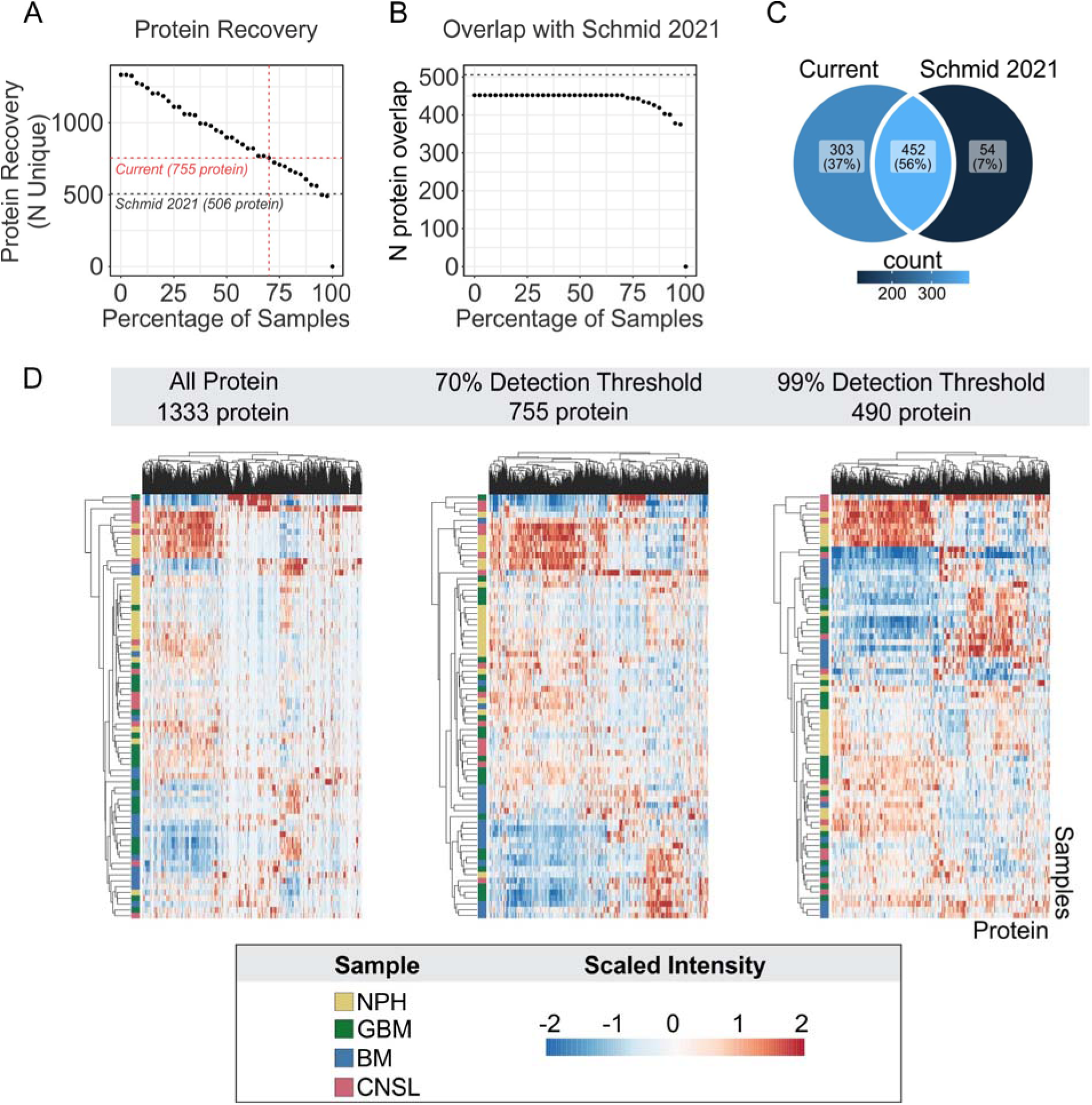
CSF protein recovery. **(A)** Relationship between recovery threshold and number of unique proteins recovered. *Dashed red line:* 70% threshold used in current study. *Dashed black line:* Number of proteins recovered by Schmid et al. (2021) for reference. **(B)** Relationship between recovery threshold and number of proteins in current study that overlap with proteins recovered by Schmid et al. *Dashed black line*: Total number of proteins recovered by Schmid et al. **(C)** Venn diagram of overlap between proteins recovered at 70% threshold in current study and Schmid et al. data. **(D)** Heatmaps illustrating scaled intensities of protein at 0% (all protein, *left*), 70% (*middle*) and 99% (*right*) recovery thresholds. **(E)** Number of unique proteins recovered stratified by diagnostic group. *p<0.05 and ***p<0.001 comparison versus NPH determined by t-test. BM; brain metastasis, GBM; glioblastoma multiforme, NPH; normal pressure hydrocephalus, CNSL; central nervous system lymphoma.

**Figure Supplement 3.**
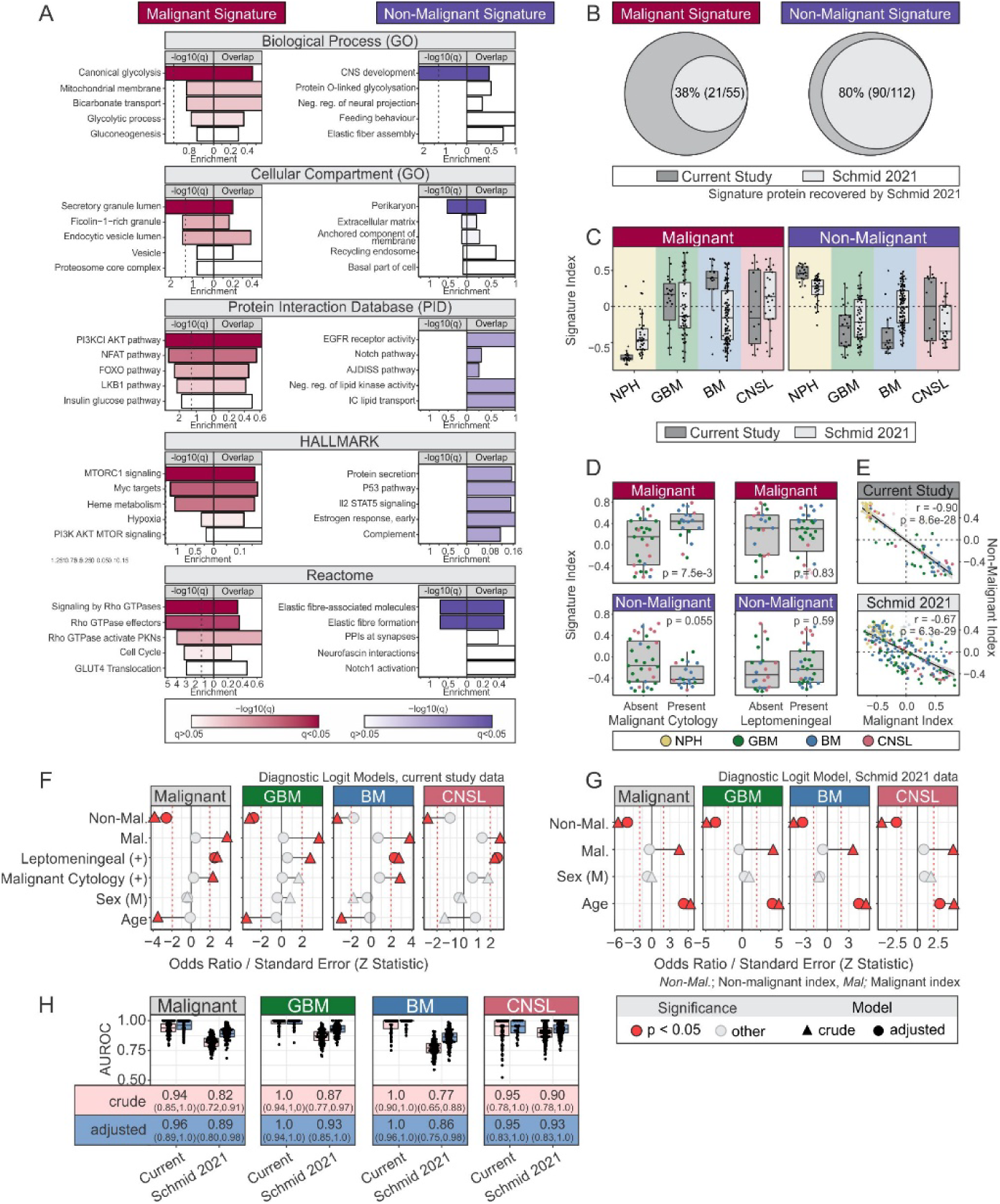
Characterization of malignant and non-malignant signatures. **(A)** Functional annotation of malignant and non-malignant signatures using gene ontology (GO), protein interaction database (PID), HALLMARK, and Reactome. *Dashed vertical line*: 5% false discovery rate (FDR) threshold, *q*: Benjamini Hochberg-adjusted p-value (i.e., FDR). **(B)** Venn diagram illustrating fraction of malignant and non-malignant signature proteins identified in current study that were recovered in in Schmid et al. (2021) CSF data. **(C)** Malignant and non-malignant indices by patient cohort. Data from current study and Schmid 2021 are shown and indices are aggregate Z scores for malignant and non-malignant signatures. **(D)** Malignant (*top*) and non-malignant indices (*bottom*) by malignant cytology presence (*left*) and leptomeningeal status (*right*). **(E)** Relationship between malignant and non-malignant indices in current study (*top*) and Schmid 2021 (*bottom*) data. **(F, G)** Forest plots of coefficients for crude (univariate) and adjusted (multivariate) LR models for current study (**F**) and Schmid 2021 (**G**). **(H)** AUROC of crude (univariate) and adjusted (multivariate) classifier models, using data from current and Schmid cohorts^16^. For F-H, adjusted models included malignant and malignant indices, leptomeningeal status, malignant cytology status, sex, and age as covariates. Only age was available as an additional covariate in the Schmid cohort.

**Figure Supplement 4.**
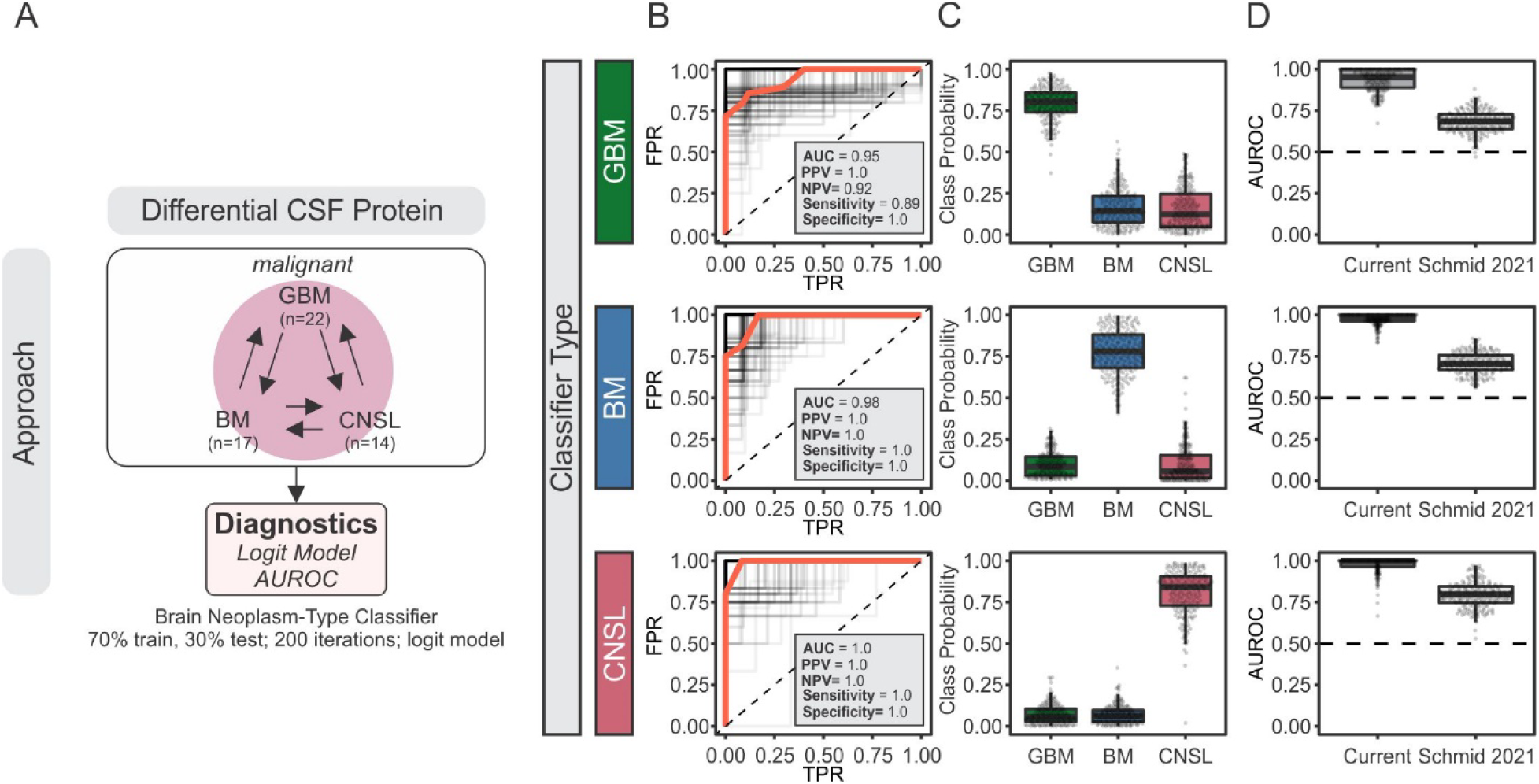
CSF proteomics discriminate between different brain neoplasms. **(A)** Schematic representation of approach to evaluate brain neoplasm discrimination by CSF proteomics. **(B-D)** Logit regression-based classifier models were trained using brain neoplasm-specific signatures and GBM (*top*), BM (*middle*) and CNSL (*bottom*) class predictions were generated. Analysis was performed using resampling procedure (200 iterations using random 70:30 train:test partitions) and evaluated with ROC curves to assess sensitivity and specificity (**B**), class probabilities (**C**) and area under ROC (AUROC) (**D**). For C, the indicated performance metrics correspond to *Model 1* in Table 2. In D, AUROCs were also evaluated in Schmid 2021 data. *Red ROC curve*: median. *Black ROC curves*: each iteration. FPR; false positive rate, TPR; true positive rate.

**Figure Supplement 5.**
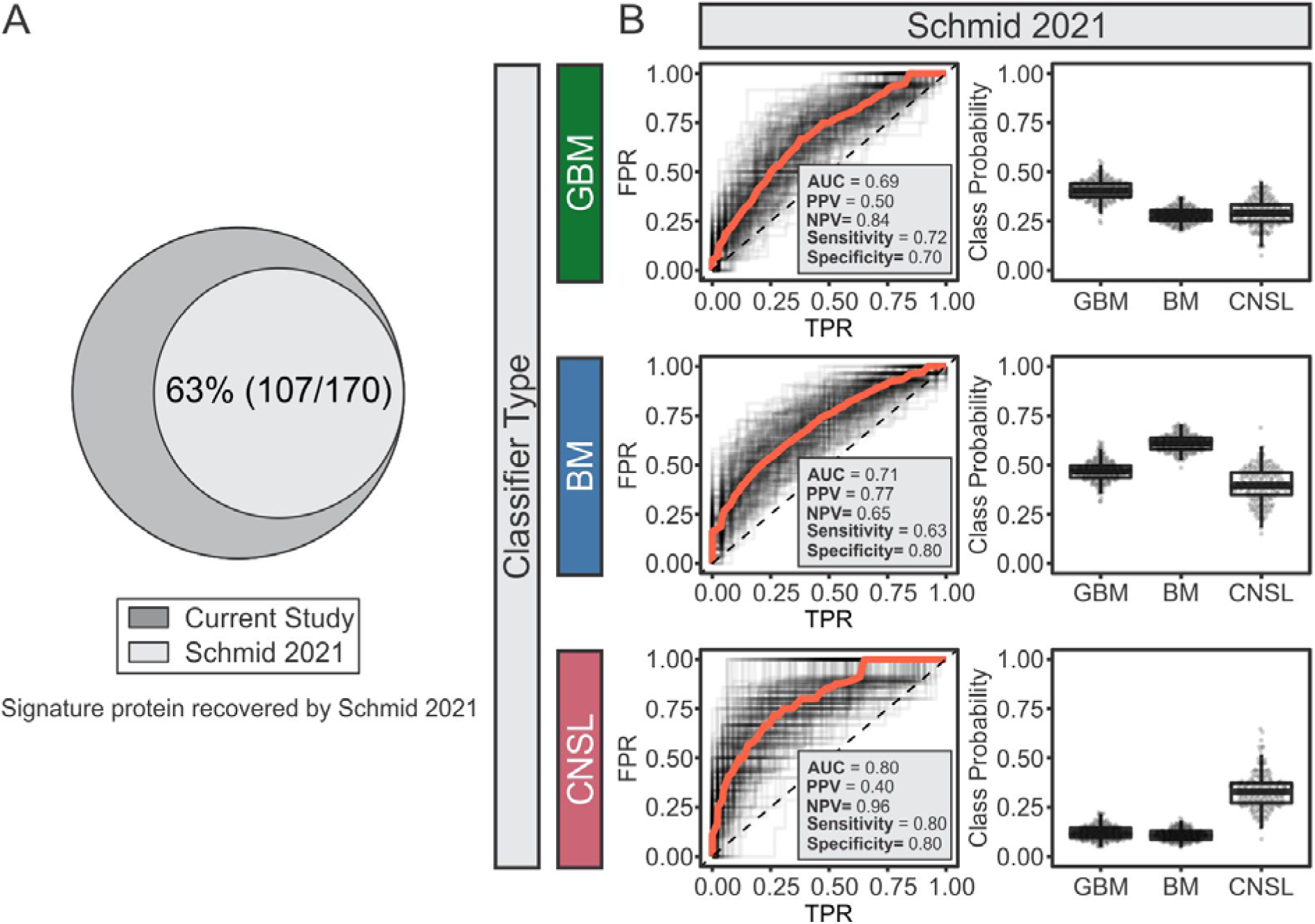
External validation of diagnostic value of brain tumor-specific biomarker signatures in CSF proteomic data from Schmid et al. (2021). **(A)** Venn diagram of neoplasm-specific biomarkers identified in current study that were recovered in in Schmid et al. (2021) CSF data. **(B)** ROC curve assessing sensitivity and specificity of brain tumor-specific proteomic signatures as diagnostic biomarkers (*left column*) and predicted class probabilities (*right column*) using Schmid et al. (2021) CSF data. The indicated performance metrics correspond to *Model 1* in Table 2. *Red ROC curve*: median. *Black ROC curves*: each iteration. FPR; false positive rate, TPR; true positive rate.

**Figure Supplement 6.**
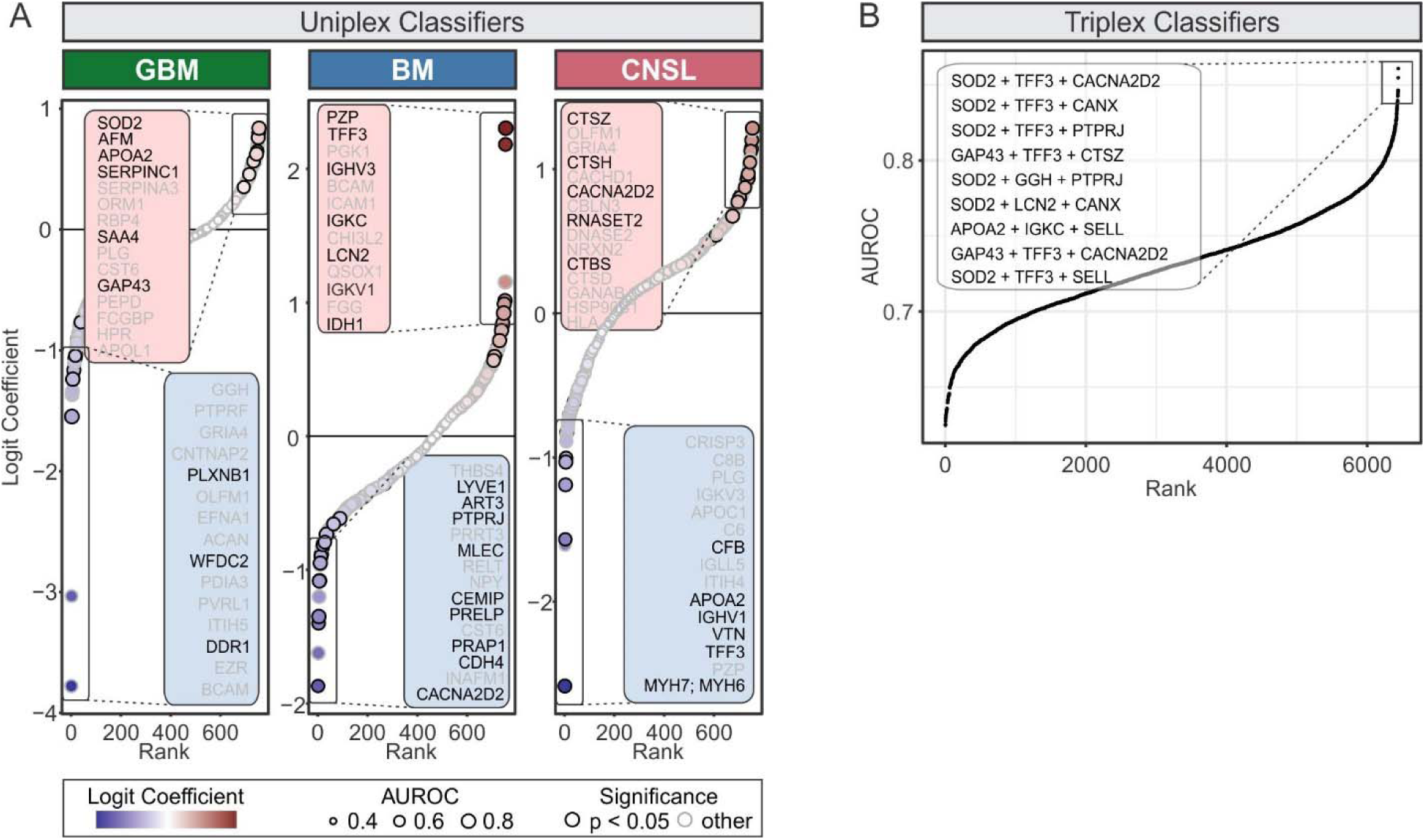
Uniplex and triplex classification models. **(A)** Candidate neoplasm-specific biomarkers identified using LR classifiers. For each protein, LR models were trained and evaluated using a resampling procedure (random 70:30 train:test partitions, 200 iterations) and these single-protein classifiers were termed uniplex classifiers. Top candidate proteins are indicated, significant hits (p < 0.05) are bolded. **(B)** AUROC plot ranking classification performance of all three-way combinations of candidate biomarkers (i.e., triplex classifier). LR; logistic regression.

**Figure Supplement 7.**
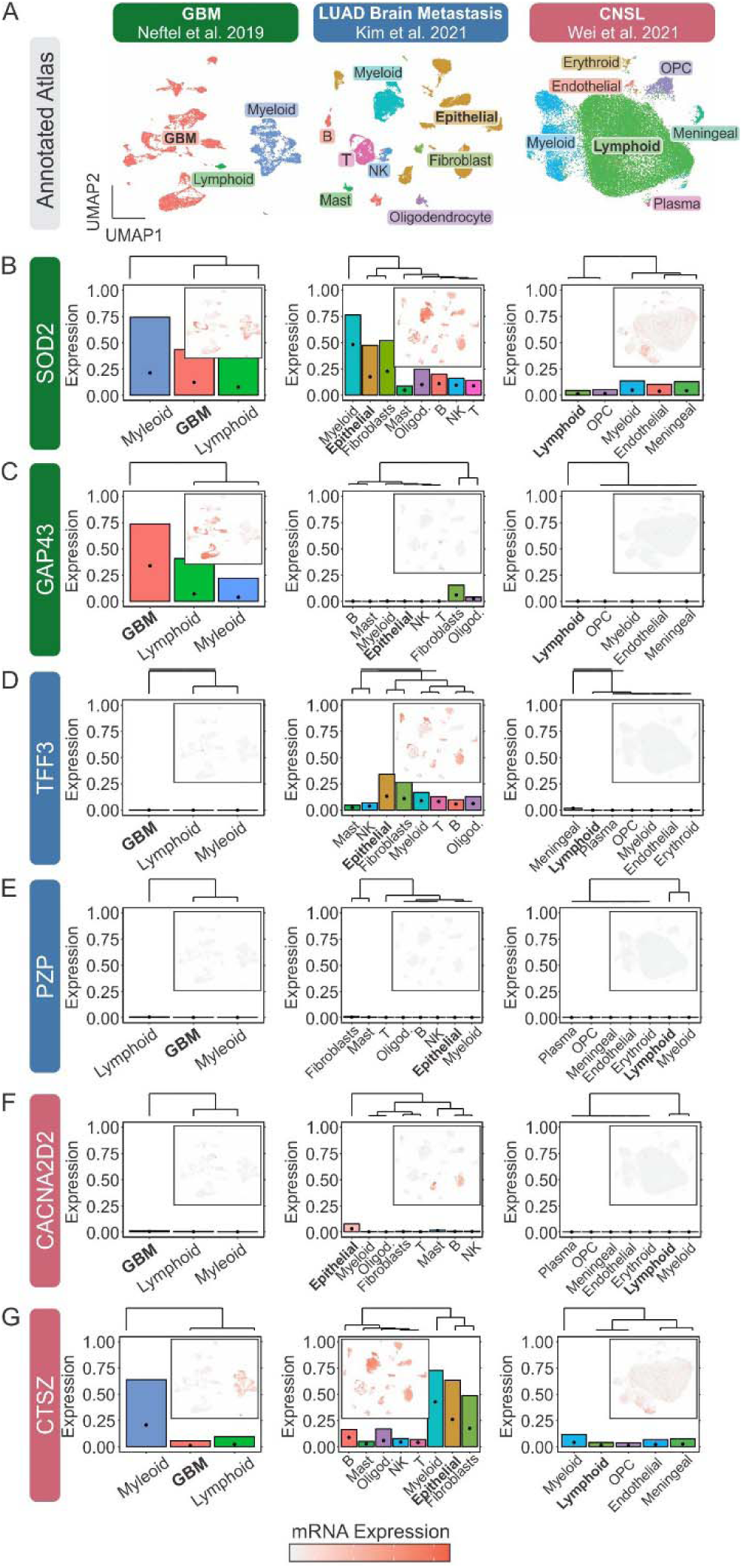
scRNAseq expression profiles of neoplasm biomarkers. (A-G) Public scRNAseq profiles of GBM (*left column*), LUAD brain metastases (*middle column*), and CNSL (*right column*) were annotated according to original reports^28–30^ (**A**) and expression of SOD2 (**B**), GAP43 (**C**), TFF3 (**D**), PZP (**E**), CACNA2D2 (**F**), and CTSZ (**G**) were visualized using UMAPs (*insets*) for cell-level profiles, and bar plots for aggregate cell-type-specific expression. *Bolded* cell labels indicate neoplastic populations. For bar plots, *bars* represent fraction of expressing cells, and *dots* represent mean normalized expression. B; B cells, GBM; glioblastoma multiforme, LUAD; lung adenocarcinoma, NK; natural kill cells, Oligod.; oligodendrocytes, OPC; oligodendrocyte progenitor cells, T; T cells.

**Figure Supplement 8.**
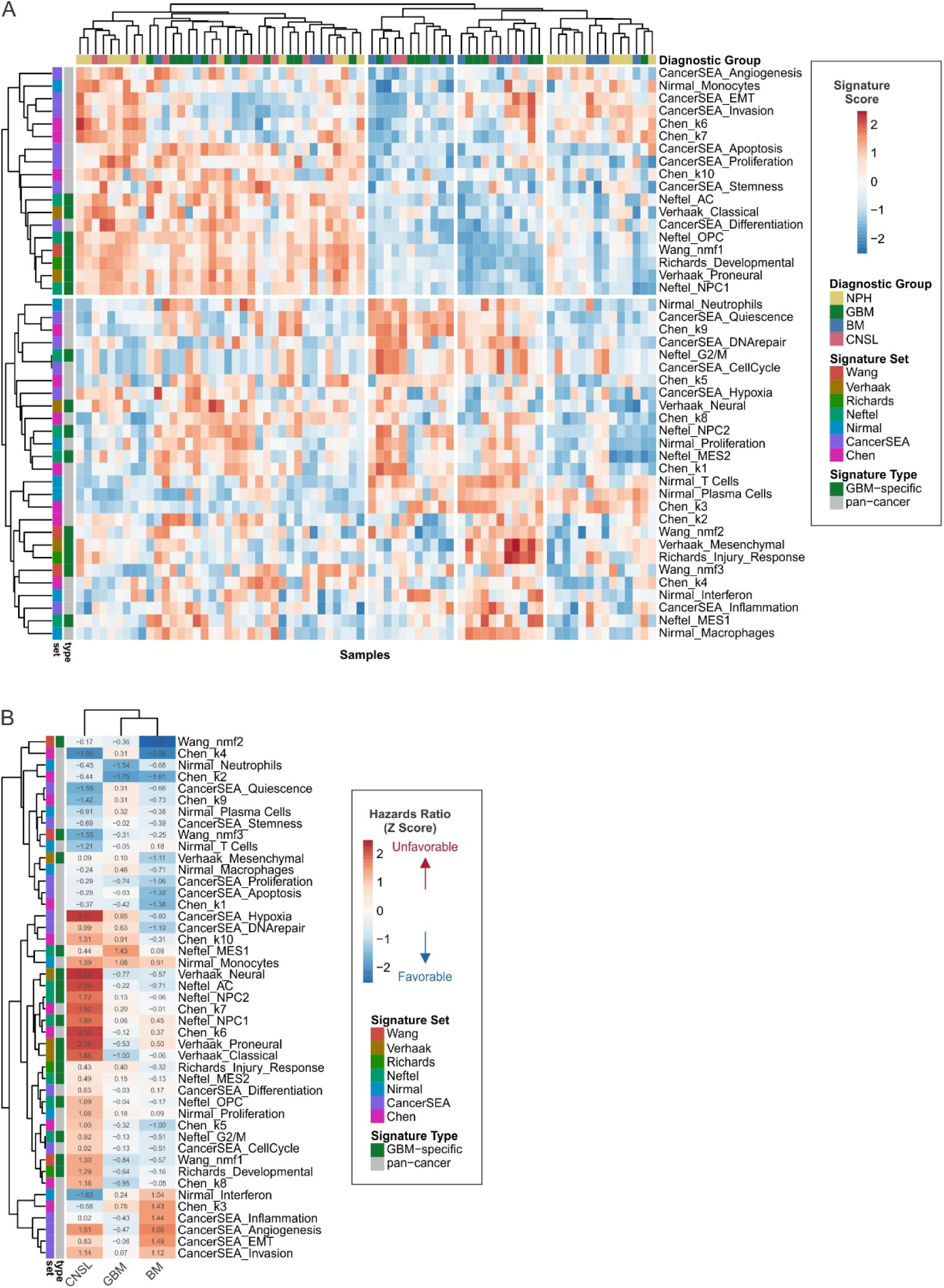
Expression and survival-associations of pan-cancer and GBM- specific signatures. **(A)** Hierarchically-clustered heatmap of signature scores. GBM-specific signature sets were obtained from Neftel et al.,^33^ Richards et al.,^32^ Verhaak et al.,^34^ and Wang et al.;^20^ whereas pan-cancer signature sets were obtained from CancerSEA,^36^ Chen et al.,^31^ and Nirmal et al.^35^ **(B)** Heatmap of hazard-ratio z scores from univariate cox proportional regression models relating signature score to survival.

**Figure Supplement 9.**
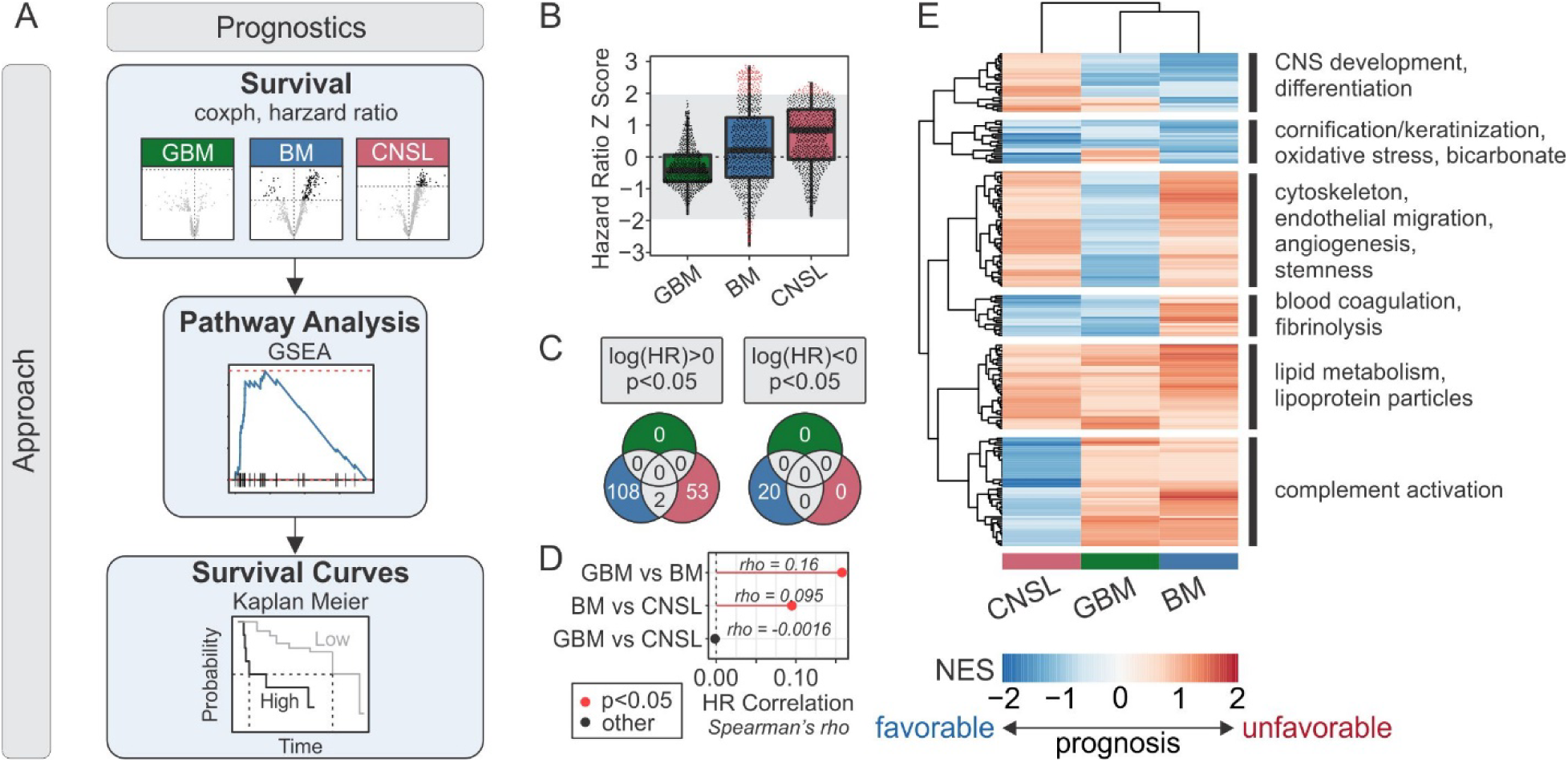
Predicting survival of brain neoplasm patients with CSF proteomics. **(A)** Schematic representation of work flow to characterize protein and pathways associated with survival. **(B)** Distribution of brain neoplasm-specific hazard ratios (HR) derived from univariate cox proportional hazards survival models for each protein. HRs are reported as Z scores, and positive and negative scores are associated with unfavorable and favorable prognosis, respectively. *Shaded grey region*: Non-significant survival models. **(C)** Venn diagram of significant prognostic biomarkers in brain neoplasms. **(D)** Spearman correlation of protein-specific hazard ratios between brain neoplasms. **(E)** Heatmap of survival-associated pathways. Gene-set enrichment analysis (GSEA) was performed on HR-ranked protein, and pathways that were significantly enriched in at least one brain neoplasm type are shown. Values in heatmap are GSEA-derived normalized enrichment scores (NES).

**Figure Supplement 10.**
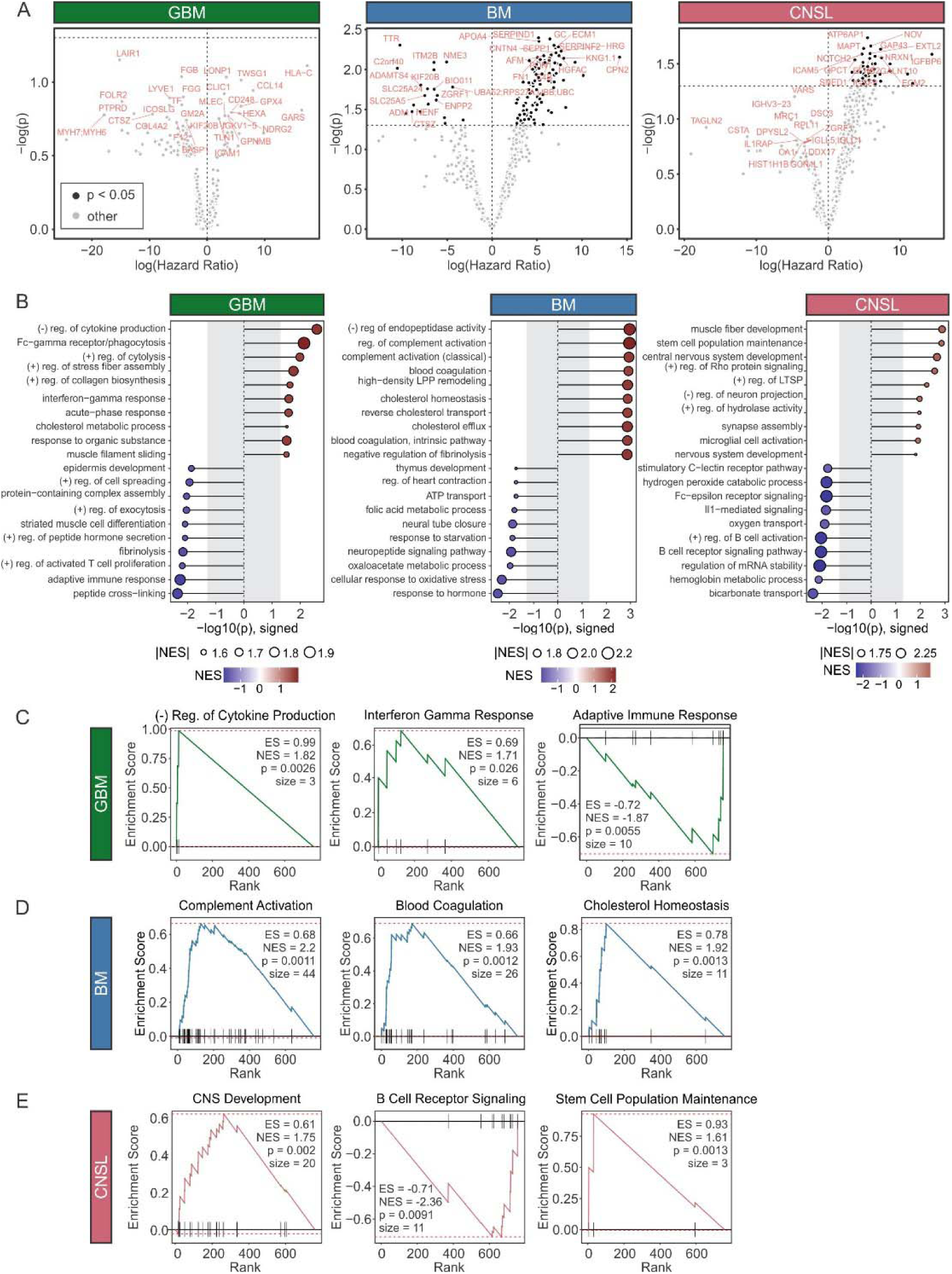
Characterization of survival-associated protein signatures. **(A)** Volcano plot of brain neoplasm-specific hazard ratios (HR) derived from univariate cox proportional hazards survival models for each protein. Positive and negative scores are associated with unfavorable and favorable prognosis, respectively, and top and bottom 15 hits are indicated in *red*. *Dashed line*: 5% significance level. **(B)** Forest plot of top GO annotations identified by GSEA performed on HR-ranked protein. **(C-E)** GSEA plots for survival-associated pathways for GBM (**C**), BM (**D**), and CNSL (**E**). ES; enrichment score, NES; normalized enrichment score.

## References

1. Ostrom, Q. T. et al. CBTRUS statistical report: primary brain and other central nervous system tumors diagnosed in the United States in 2012–2016. Neuro-oncology 21, v1–v100 (2019).

2. Stupp, R. et al. Effects of radiotherapy with concomitant and adjuvant temozolomide versus radiotherapy alone on survival in glioblastoma in a randomised phase III study: 5-year analysis of the EORTC-NCIC trial. The lancet oncology 10, 459–466 (2009).

3. Stupp, R. et al. Effect of Tumor-Treating Fields Plus Maintenance Temozolomide vs Maintenance Temozolomide Alone on Survival in Patients With Glioblastoma: A Randomized Clinical Trial. JAMA 318, 2306–2316, doi:10.1001/jama.2017.18718 (2017).

4. O’Connell, K., Romo, C. G. & Grossman, S. A. Brain metastases as a first site of recurrence in patients receiving chemotherapy with controlled systemic cancer: a critical but under-recognized clinical scenario. Current treatment options in neurology 21, 1–7 (2019).

5. Nieder, C., Spanne, O., Mehta, M. P., Grosu, A. L. & Geinitz, H. Presentation, patterns of care, and survival in patients with brain metastases: what has changed in the last 20 years? Cancer 117, 2505–2512 (2011).

6. Andre, F. et al. Patterns of relapse of N2 nonsmall cell lung carcinoma patients treated with preoperative chemotherapy: Should prophylactic cranial irradiation be reconsidered? Cancer 91, 2394–2400 (2001).

7. Freilich, R. J., Seidman, A. D. & Deangelis, L. M. Central nervous system progression of metastatic breast cancer in patients treated with paclitaxel. Cancer 76, 232–236 (1995).

8. Malone, H. et al. Complications following stereotactic needle biopsy of intracranial tumors. World neurosurgery 84, 1084–1089 (2015).

9. Kucharczyk, M. J., Parpia, S., Whitton, A. & Greenspoon, J. N. Evaluation of pseudoprogression in patients with glioblastoma. Neuro-oncology practice 4, 120–134 (2017).

10. Zikou, A. et al. Radiation necrosis, pseudoprogression, pseudoresponse, and tumor recurrence: imaging challenges for the evaluation of treated gliomas. Contrast media & molecular imaging 2018 (2018).

11. Miller, A. M. et al. Tracking tumour evolution in glioma through liquid biopsies of cerebrospinal fluid. Nature 565, 654–658 (2019).

12. De Mattos-Arruda, L. et al. Cerebrospinal fluid-derived circulating tumour DNA better represents the genomic alterations of brain tumours than plasma. Nature communications 6 (2015).

13. Newman, A. M. et al. An ultrasensitive method for quantitating circulating tumor DNA with broad patient coverage. Nature medicine 20, 548–554 (2014).

14. Nassiri, F. et al. Detection and discrimination of intracranial tumors using plasma cell-free DNA methylomes. Nature Medicine 26, 1044–1047 (2020).

15. Shen, S. Y. et al. Sensitive tumour detection and classification using plasma cell-free DNA methylomes. Nature 563, 579–583 (2018).

16. Schmid, D. et al. Diagnostic biomarkers from proteomic characterization of cerebrospinal fluid in patients with brain malignancies. Journal of neurochemistry (2021).

17. Zhang, H. et al. Integrated proteogenomic characterization of human high-grade serous ovarian cancer. Cell 166, 755–765 (2016).

18. Mertins, P. et al. Proteogenomics connects somatic mutations to signalling in breast cancer. Nature 534, 55–62 (2016).

19. Oh, S. et al. Integrated pharmaco-proteogenomics defines two subgroups in isocitrate dehydrogenase wild-type glioblastoma with prognostic and therapeutic opportunities. Nature communications 11, 1–16 (2020).

20. Wang, L. B. et al. Proteogenomic and metabolomic characterization of human glioblastoma. Cancer Cell 39, 509–528.e520, doi:10.1016/j.ccell.2021.01.006 (2021).

21. Bailey, M. H. et al. Comprehensive characterization of cancer driver genes and mutations. Cell 173, 371–385. e318 (2018).

22. Bader, J. M. et al. Proteome profiling in cerebrospinal fluid reveals novel biomarkers of Alzheimer’s disease. Molecular systems biology 16, e9356 (2020).

23. Bereman, M. S., Beri, J., Enders, J. R. & Nash, T. Machine learning reveals protein signatures in CSF and plasma fluids of clinical value for ALS. Scientific reports 8, 1–14 (2018).

24. Stoop, M. P. et al. Decreased NeuroLJAxonal Proteins in CSF at First Attack of Suspected Multiple Sclerosis. PROTEOMICS–Clinical Applications 11, 1700005 (2017).

25. Sinha, A. et al. N-Glycoproteomics of patient-derived xenografts: a strategy to discover tumor-associated proteins in high-grade serous ovarian cancer. Cell Systems 8, 345–351. e344 (2019).

26. Kim, Y. et al. Targeted proteomics identifies liquid-biopsy signatures for extracapsular prostate cancer. Nature Communications 7, 11906, doi:10.1038/ncomms11906 (2016).

27. Guldbrandsen, A. et al. Development of robust targeted proteomics assays for cerebrospinal fluid biomarkers in multiple sclerosis. Clinical proteomics 17, 1–21 (2020).

28. Neftel, C. et al. An integrative model of cellular states, plasticity, and genetics for glioblastoma. Cell 178, 835–849. e821 (2019).

29. Kim, N. et al. Single-cell RNA sequencing demonstrates the molecular and cellular reprogramming of metastatic lung adenocarcinoma. Nature communications 11, 1–15 (2020).

30. Wei, B. et al. Analysis of Cellular Heterogeneity in Immune Microenvironment of Primary Central Nervous System Lymphoma by Single-Cell Sequencing. Frontiers in oncology 11 (2021).

31. Chen, F., Chandrashekar, D. S., Varambally, S. & Creighton, C. J. Pan-cancer molecular subtypes revealed by mass-spectrometry-based proteomic characterization of more than 500 human cancers. Nature Communications 10, 5679, doi:10.1038/s41467-019-13528-0 (2019).

32. Richards, L. M. et al. Gradient of Developmental and Injury Response transcriptional states defines functional vulnerabilities underpinning glioblastoma heterogeneity. Nature Cancer 2, 157–173, doi:10.1038/s43018-020-00154-9 (2021).

33. Neftel, C. et al. An Integrative Model of Cellular States, Plasticity, and Genetics for Glioblastoma. Cell 178, 835–849.e821, doi:10.1016/j.cell.2019.06.024 (2019).

34. Verhaak, R. G. W. et al. Integrated genomic analysis identifies clinically relevant subtypes of glioblastoma characterized by abnormalities in PDGFRA, IDH1, EGFR, and NF1. Cancer cell 17, 98–110, doi:10.1016/j.ccr.2009.12.020 (2010).

35. Nirmal, A. J. et al. Immune Cell Gene Signatures for Profiling the Microenvironment of Solid Tumors. Cancer Immunol Res 6, 1388–1400, doi:10.1158/2326-6066.Cir-18-0342 (2018).

36. Yuan, H. et al. CancerSEA: a cancer single-cell state atlas. Nucleic Acids Res 47, D900–d908, doi:10.1093/nar/gky939 (2019).

37. Potriquet, J., Laohaviroj, M., Bethony, J. M. & Mulvenna, J. A modified FASP protocol for high-throughput preparation of protein samples for mass spectrometry. PloS one 12, e0175967 (2017).

38. Ronsein, G. E. et al. Parallel reaction monitoring (PRM) and selected reaction monitoring (SRM) exhibit comparable linearity, dynamic range and precision for targeted quantitative HDL proteomics. Journal of proteomics 113, 388–399 (2015).

39. Mao, J. et al. Convenient multicomponent reaction synthesis of novel pyrano [4, 3-b] pyran derivatives via a domino reaction under microwave irradiation. Arkivoc 3, 171–186 (2016).

40. Brown, C. E. et al. Regression of glioblastoma after chimeric antigen receptor T-cell therapy. New England Journal of Medicine 375, 2561–2569 (2016).

41. Samson, M. H. et al. Trefoil factor family peptides in human saliva and cyclical cervical mucus. Method evaluation and results on healthy individuals. Clinical chemistry and laboratory medicine 49, 861–868 (2011).

42. Hoffmann, W. Trefoil factors. Cellular and Molecular Life Sciences CMLS 62, 2932–2938 (2005).

43. Chen, Y.-H., Lu, Y., De Plaen, I. G., Wang, L.-Y. & Tan, X.-D. Transcription factor NF-κB signals antianoikic function of trefoil factor 3 on intestinal epithelial cells. Biochemical and biophysical research communications 274, 576–582 (2000).

44. Kinoshita, K., Taupin, D. R., Itoh, H. & Podolsky, D. K. Distinct pathways of cell migration and antiapoptotic response to epithelial injury: structure-function analysis of human intestinal trefoil factor. Molecular and cellular biology 20, 4680–4690 (2000).

45. Smid, M. et al. Genes associated with breast cancer metastatic to bone. Journal of Clinical Oncology 24, 2261–2267 (2006).

46. Ahmed, A. R., Griffiths, A. B., Tilby, M. T., Westley, B. R. & May, F. E. TFF3 is a normal breast epithelial protein and is associated with differentiated phenotype in early breast cancer but predisposes to invasion and metastasis in advanced disease. The American journal of pathology 180, 904–916 (2012).

47. Magbanua, M. J. M. et al. Molecular profiling of tumor cells in cerebrospinal fluid and matched primary tumors from metastatic breast cancer patients with leptomeningeal carcinomatosis. Cancer research 73, 7134–7143 (2013).

48. Taupin, D. et al. Augmented intestinal trefoil factor (TFF3) and loss of pS2 (TFF1) expression precedes metaplastic differentiation of gastric epithelium. Laboratory investigation 81, 397–408 (2001).

49. Terris, B. et al. Characterization of gene expression profiles in intraductal papillary-mucinous tumors of the pancreas. The American journal of pathology 160, 1745–1754 (2002).

50. Taupin, D., Ooi, K., Yeomans, N. & Giraud, A. Conserved expression of intestinal trefoil factor in the human colonic adenoma-carcinoma sequence. Laboratory investigation; a journal of technical methods and pathology 75, 25–32 (1996).

51. Yuan, Z., Chen, D., Chen, X., Yang, H. & Wei, Y. Overexpression of trefoil factor 3 (TFF3) contributes to the malignant progression in cervical cancer cells. Cancer cell international 17, 1–13 (2017).

52. Faith, D. A. et al. Trefoil factor 3 overexpression in prostatic carcinoma: prognostic importance using tissue microarrays. The Prostate 61, 215–227 (2004).

53. Achrol, A. S. et al. Brain metastases. Nature Reviews Disease Primers 5, 1–26 (2019).

54. Sperduto, P. W. et al. The effect of gene alterations and tyrosine kinase inhibition on survival and cause of death in patients with adenocarcinoma of the lung and brain metastases. International Journal of Radiation Oncology* Biology* Physics 96, 406–413 (2016).

55. Zimmer, A. S., Van Swearingen, A. E. & Anders, C. K. HER2LJpositive breast cancer brain metastasis: A new and exciting landscape. Cancer Reports, e1274 (2020).

56. Fiske, J. L., Fomin, V. P., Brown, M. L., Duncan, R. L. & Sikes, R. A. Voltage-sensitive ion channels and cancer. Cancer and Metastasis Reviews 25, 493–500 (2006).

57. Kunzelmann, K. Ion channels and cancer. The Journal of membrane biology 205, 159–173 (2005).

58. Lerman, M. I. & Minna, J. D. The 630-kb lung cancer homozygous deletion region on human chromosome 3p21. 3: identification and evaluation of the resident candidate tumor suppressor genes. Cancer research 60, 6116–6133 (2000).

59. Gao, B. et al. Functional properties of a new voltage-dependent calcium channel α2δ auxiliary subunit gene (CACNA2D2). Journal of Biological Chemistry 275, 12237–12242 (2000).

60. Zhang, Y., Liu, Y., Liu, H. & Tang, W. H. Exosomes: biogenesis, biologic function and clinical potential. Cell & Bioscience 9, 19, doi:10.1186/s13578-019-0282-2 (2019).

61. Hornung, S., Dutta, S. & Bitan, G. CNS-Derived Blood Exosomes as a Promising Source of Biomarkers: Opportunities and Challenges. Frontiers in Molecular Neuroscience 13, doi:10.3389/fnmol.2020.00038 (2020).

62. Berger, S. T. et al. MStern Blotting-High Throughput Polyvinylidene Fluoride (PVDF) Membrane-Based Proteomic Sample Preparation for 96-Well Plates. Mol Cell Proteomics 14, 2814–2823, doi:10.1074/mcp.O115.049650 (2015).

63. Cox, J. & Mann, M. MaxQuant enables high peptide identification rates, individualized p.p.b.-range mass accuracies and proteome-wide protein quantification. Nature Biotechnology 26, 1367–1372, doi:10.1038/nbt.1511 (2008).

64. Wojtowicz, E. E. et al. Ectopic miR-125a Expression Induces Long-Term Repopulating Stem Cell Capacity in Mouse and Human Hematopoietic Progenitors. Cell Stem Cell 19, 383–396, doi:10.1016/j.stem.2016.06.008 (2016).

65. Hao, Y. et al. Integrated analysis of multimodal single-cell data. Cell (2021).

66. Stuart, T. et al. Comprehensive integration of single-cell data. Cell 177, 1888–1902. e1821 (2019).

67. Butler, A., Hoffman, P., Smibert, P., Papalexi, E. & Satija, R. Integrating single-cell transcriptomic data across different conditions, technologies, and species. Nature biotechnology 36, 411–420 (2018).

68. Satija, R., Farrell, J. A., Gennert, D., Schier, A. F. & Regev, A. Spatial reconstruction of single-cell gene expression data. Nature biotechnology 33, 495–502 (2015).

69. Todorov, V. & Todorov, M. V. Package ‘rrcov’. (2016).

70. Korsunsky, I., Nathan, A., Millard, N. & Raychaudhuri, S. Presto scales Wilcoxon and auROC analyses to millions of observations. BioRxiv, 653253 (2019).

71. Korotkevich, G. et al. Fast gene set enrichment analysis. BioRxiv, 060012 (2021).

72. Liberzon, A. et al. The molecular signatures database hallmark gene set collection. Cell systems 1, 417–425 (2015).

73. Fabregat, A. et al. The reactome pathway knowledgebase. Nucleic acids research 46, D649–D655 (2018).

74. Gelman, A. et al. Package ‘arm’. Data Analysis Using Regression and Multilevel/Hierarchical Models (2015).

75. Sing, T., Sander, O., Beerenwinkel, N. & Lengauer, T. ROCR: visualizing classifier performance in R. Bioinformatics 21, 3940–3941 (2005).

76. Hänzelmann, S., Castelo, R. & Guinney, J. GSVA: gene set variation analysis for microarray and RNA-seq data. BMC bioinformatics 14, 1–15 (2013).

77. Kassambara, A., Kosinski, M., Biecek, P. & Fabian, S. Package ‘survminer’. Drawing Survival Curves using ‘ggplot2’(R package version 03 1) (2017).

